# Educational attainment of children with major congenital anomalies during primary school in England: a population cohort study using linked administrative data from ECHILD

**DOI:** 10.1101/2025.06.21.25329922

**Authors:** Joachim Tan, Ayana Cant, Kate Lewis, Vincent Nguyen, Laura Gimeno, Ania Zylbersztejn, Pia Hardelid, Joan Morris, Bianca De Stavola, Katie Harron, Ruth Gilbert, the HOPE Study Team

**Author notes:** Corresponding author Joachim Tan UCL Great Ormond Street Institute of Child Health University College London London WC1N 1EH United Kingdom.

## Abstract

**Background:** Major congenital anomalies (CAs) occur in 2.3% of livebirths and are associated with lower educational attainment in affected children. Understanding how attainment changes throughout primary school would inform parents, schools and organisations and help plan support.

**Objectives:** We compared school enrolment and attainment at ages 5, 7 and 11 in children with different CAs and their peers in England using linked administrative hospital and education data in the ECHILD database.

**Methods:** We included all singleton children born in NHS-funded hospitals from September 2003 to August 2008 who enrolled in state-funded schools at age 4-5. CAs were identified from hospital diagnoses, procedures or death records. We described school enrolment, school-readiness, the percentages of children who sat curriculum assessments and who achieved expected levels in English and Maths at three ages. We estimated risk ratios of children with CAs achieving expected levels compared with peers, adjusting for sociodemographic factors.

**Results:** Of 2,351,589 singleton children enrolled at age 5, 78,847 (3.5%) had CAs. At age 11, 88.7% of enrolled children with CAs sat assessments versus 97.2% of peers. Proportionally fewer children with CAs (45.7%) were school-ready at age 5 versus peers (57.0%). For English, 56.9%, 55.4% and 65.3% of children with CAs achieved expected levels at ages 5, 7 and 11 respectively, consistently 11%-12% fewer than peers; similar gaps persisted for Maths. Children with CAs were on average less likely than peers to achieve expected levels [adjusted risk ratio, aRR (95%CI): 0.86 (0.85,0.86)] but this varied substantially across CA subgroups [aRR (95%CI) range: 0.01 (0.01,0.02) to 1.04 (0.96,1.12)].

**Conclusion:** The attainment gap between children with CAs and peers remained unchanged across subjects and ages, with proportionally fewer sitting assessments at age 11. Better monitoring and support for these children from school entry could help optimise learning experiences and fulfil their academic potential.

**SYNOPSIS:** *Study question:* What are the patterns of educational attainment in children with major congenital anomalies (CAs) throughout primary school?

*What is already known:* Studies of regional or registry-specific cohorts of children with CAs showed that proportionally fewer achieve expected attainment relative to peers at single stages of national assessments. There is limited evidence on children’s participation in assessments and how attainment gaps change throughout primary school at a population level.

*What the study adds:* Longitudinal analysis of whole-population cohorts from ages 4 to 11 found that attainment gaps between children with and without CAs remained largely constant across ages. Over half of children with CAs were assessed as not ’school-ready’ at age 5. Whilst almost all children with CAs remained enrolled in school at age 11, one in nine did not participate in assessments.

## 1 | BACKGROUND

Major congenital anomalies (hereafter CAs), are inborn structural, chromosomal or genetic disorders with significant medical, functional or social consequences for individuals. Many CAs are rare diseases, affecting under 1 in 2,000 people, but collectively they occur in 2.3% of births in England.^1^ Progressively improving survival of children with CAs have led to more of them starting school,^2^ but fewer achieve expected levels of attainment compared with their peers.^3–5^ Complex health needs, higher rates of school absences, inherent learning disabilities, and inadequate support for special educational needs, are likely contributors to adverse outcomes for affected children, including those with non-chromosomal CAs such as cardiac defects, orofacial clefts or spina bifida.^6–8^ Such findings were corroborated by a recent study using linked education and regional CA registries’ data in England, which showed that attainment rates in national tests at ages 11 and 16 for children with a range of structural CAs were on average 5-7% lower than their peers.^9^

This study aimed to describe the attainment trajectories for children with and without CAs in England by following whole-population cohorts through primary school (ages 4-11 years). Understanding how the rates of school enrolment, curriculum assessments and attainment in children with different CAs change from school entry will provide evidence to inform timely support, thereby enriching their educational experience and maximising their academic potential during these formative years. This project contributes to the Health Outcomes of young People in Education (HOPE) research programme, previously described elsewhere.^10^

## 2 | METHODS

### 2.1 | Study Design

This is a population-based retrospective cohort study using linked administrative data from the ECHILD (Education and Child Health Insights from Linked Data) database.^11^

#### 2.1.1 | Data Sources

ECHILD contains routinely-collected data on hospital admissions from Hospital Episode Statistics (HES) linked to education data from the National Pupil Database (NPD). HES captures about 97% of births in NHS-funded hospitals in England and 98-99% of secondary care contacts, including information on demographic and geographical details, hospital stays, diagnoses and procedures^12^; the version of ECHILD used covered approximately 14.7 million individuals born 1995-2020.^10, 11^ ECHILD also contains death registrations and causes of death from the Office for National Statistics. Diagnoses and causes of death are coded using the International Classification of Diseases 10^th^ Revision (ICD-10) whilst procedures are coded using the Office of Population Censuses and Surveys Classification of Interventions and Procedures 4th Revision (OPCS-4). The NPD holds information on children attending state-funded schools in England between the ages of 4-18 years from academic year 2001/02.^13^ Data include teacher-assessed outcomes and national test marks at different key stages of education, pupils’ ethnicity, area-based deprivation indices, free school meals eligibility (FSME) and geographical information.

#### 2.1.2 | Cohort Selection

The study population comprised all singleton children born in NHS-funded hospitals between 1st September 2003 and 31st August 2008 who were linked to the NPD. We included children who were enrolled in Reception year (age 4/5) in state-funded schools, as recorded in the January school census. We excluded children who were two or more years outside of the expected age for their school year. Children were followed up until end of primary school (age 11), enrolment ceased or death (whichever was earlier).

### 2.2 | Major congenital anomalies

CA subgroups were defined by ICD-10 diagnosis codes according to the EUROCAT (European network of population-based registries for the surveillance of congenital anomalies) guide version 1.4.^14^ We also used alternative codelists combining diagnosis and OPCS-4 codes for severe congenital heart defects (CHD),^15^ orofacial clefts,^3, 16, 17^ anorectal malformations^18^ and hypospadias^19^; all except severe CHD produced more conservative birth prevalence estimates than EUROCAT. We searched for relevant diagnosis codes recorded before the first birthday and procedure codes (where specified) in HES, or causes of death up to the 12th birthday.

We described results for exemplar cardiac, orofacial, digestive, renal, and limb CAs and syndromic Cas (e.g. Down, Turner, Di George syndromes), and specific CAs where NPD data had been used.^3, 4^ For non-syndromic CAs, we focused on the subset of children with isolated anomalies – i.e. one or more structural defects occurring in the same organ system (e.g. polydactyly of hands and feet) classified according to a EUROCAT algorithm.^20^ Those with CAs in multiple organ systems (not linked by a known sequence or chromosomal/genetic malformation) from a heterogeneous group and served as comparators in sensitivity analysis (see below). CA subgroups are described in eTable1.

### 2.3 | Outcomes

In England, academic assessments based on the National Curriculum are conducted at the end of three key stages of primary school – Early Years Foundation Stage Profile (EYFSP), Key Stage 1 (KS1) and Key Stage 2 (KS2) – interchangeably referred to using ages 5, 7 and 11 respectively. Specific to EYFSP is whether children reach a Good Level of Development (GLD), a composite indicator of school readiness.^21^ Attainment in English and Maths are measured by whether children achieve the age-specific expected level, based on teacher-assessments (EYFSP and KS1) and nationally marked tests (KS2). Last, continuous subject z-scores (transformed from attained levels or test marks and standardised using raw scores from all pupils assessed in an academic year) were used to compare attainment across key stages. Details of assessment metrics are in eTable2.

### 2.4 | Covariates

We presented birth and sociodemographic characteristics including potential confounders of the association between CA status and outcomes: sex at birth, academic year of birth, month of birth, maternal age at birth, birthweight, gestational age, ethnicity, income deprivation affecting children index (IDACI) quintile, free school meals eligibility (FSME) and region of pupils’ residence. The first six are contained in HES whilst the remainder are from the NPD.

### 2.5 | Statistical Methods

We quantified the number of children who were enrolled in the Spring Census at ages 5, 7, and 11, and those who died during primary school (with % relative to those enrolled at age 5). Using all enrolled children as the denominator, we reported the percentage: (1) who reached GLD; (2) who were assessed; (3) who achieved expected levels; and (4) the distribution of z-scores, for each subject and key stage, by CA subgroup.

Generalised linear models (Poisson distribution, log link and robust standard errors) were used to estimate risk ratios of achieving expected levels for English and Maths at EYFSP, KS1 and KS2, comparing children in CA subgroups to children without CAs. We coded both “not assessed” and “assessed but not achieved” as “not achieved”. Models were adjusted for sex, and then for sex and maternal age, ethnicity, IDACI quintile and FSME; year/month of birth and region were not significant confounders, whilst birthweight and gestational age were likely mediators.^22, 23^

For selected CA subgroups and all CAs combined, we fitted linear mixed models with random intercepts and random slopes to estimate the trajectories of z-scores, comparing children with and without CAs, adjusting for the factors described above. We included interaction terms to allow the differentials by CA status and sex to vary with age. To reduce computation time, a random sample consisting of 25% of children without CAs were used as the reference group. All analyses were performed using Stata version 18 (StataCorp LLC, Texas, USA).

#### 2.5.1 | Missing Data

Sex at birth, if missing, was substituted with gender recorded in the NPD. For region, FSME and IDACI, if data were missing at age 5, we used the earliest non-missing record in any subsequent school census. Ethnicity was based on the modal value across all censuses. Children in census without assessment results were considered as not achieving the expected level. For all regression models, we analysed complete cases as <5% of the total had any missing covariate value.

#### 2.5.2 | Sensitivity Analysis

We compared outcomes for CA subgroups defined by EUROCAT and alternative codelists. We stratified attainment results by malformation type (isolated, multiple or chromosomal/genetic) to check that our results were consistent with expectations of better attainment in the first group. We estimated risk ratios using two populations: (1) all children enrolled at a given key stage; (2) a subset who were assessed at all three key stages; smaller differences were expected between the latter group and peers.

### 2.6 | Ethics

Permissions to use linked, de-identified data from Hospital Episode Statistics and the National Pupil Database were granted by the Department of Education (DR200604.02B) and NHS Digital (DARS-NIC-381972). Ethical approval for the ECHILD project was granted by the National Research Ethics Service (17/LO/1494), NHS Health Research Authority Research Ethics Committee (20/EE/0180 and 21/SW/0159), and UCL Great Ormond Street Institute of Child Health’s Joint Research and Development Office (20PE06).

## 3 | RESULTS

A total of 3,042,909 livebirths were extracted from ECHILD. Of the 2,940,072 (96.6%) singleton children who were alive at their 4th birthday, 421,066 (13.8%) could not be linked to the NPD, whilst 167,417 (5.5%) who linked to the NPD were not enrolled at age 5, leaving just over 2.35 million children in our study (Figure 1).

**Figure 1:**
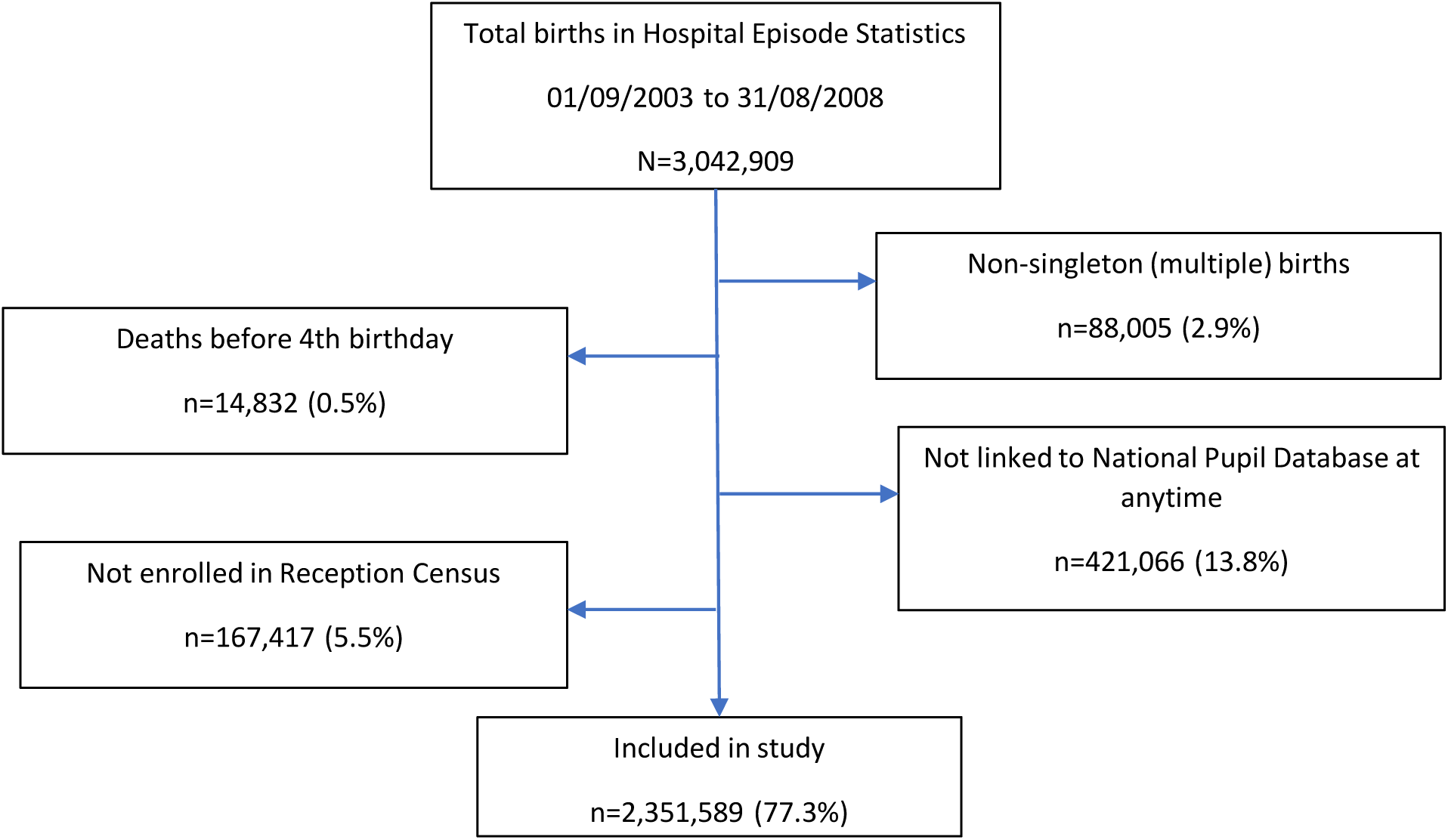
Flowchart showing starting population, exclusions and final numbers included in study

Of the study population, there were 78,847 (3.5%) children with one or more CA. Among them 71.5% had isolated CAs, another 20.3% had chromosomal, genetic or non-system specific CAs, whilst 8.2% had potential multiple CAs (Table 1). Of all children without CAs enrolled at age 5, 1.4% and 4.0% were not subsequently enrolled at ages 7 and 11 respectively, whilst corresponding percentages were 1.3% and 3.9% for children with CAs. Mortality during primary school was 0.4% in children with CAs, 10-fold higher than peers (eTable3).

**Table 1:** Birth prevalence and number included in study, by congenital anomaly (CA) subgroup and malformation type

| subgroup | Number of cases in HES | Birth prevalence (per 10,000) | Enrolled in Reception | MALFORMATION TYPE<sup>a</sup> |  |  |
| --- | --- | --- | --- | --- | --- | --- |
|  |  |  |  | Isolated | Potential multiple | Genetic, chromosomal or other |
| **Any CA** | 105,514 | 346.8 | 78,847 | 71.5% | 8.2% | 20.3% |
| Neural Tube Defects | 833 | 2.7 | 510 | 53.9% | 42.5% | 3.5% |
| Hydrocephalus | 1,180 | 3.9 | 701 | 55.6% | 39.2% | 5.1% |
| Congenital Cataract | 614 | 2.0 | 470 | 81.3% | 11.5% | 7.2% |
| **Congenital Heart Defects (CHD)** | 23,792 | 78.2 | 15,852 | 71.7% | 16.1% | 12.2% |
| Ventricular Septal Defect without severe CHD | 5,075 | 16.7 | 3,673 | 71.6% | 14.6% | 13.8% |
| Pulmonary Valve Stenosis without severe CHD | 702 | 2.3 | 546 | 78.6% | 11.9% | 9.5% |
| PDA as only CHD in term infants | 4,442 | 14.6 | 2,611 | 79.2% | 15.3% | 5.5% |
| **Severe CHD** | 6,630 | 21.8 | 4,283 | 69.8% | 14.0% | 16.2% |
| Atrioventricular Septal Defect | 1,398 | 4.6 | 832 | 40.5% | 11.5% | 48.0% |
| Tetralogy of Fallot | 1,162 | 3.8 | 826 | 63.9% | 17.9% | 18.2% |
| Hypoplastic Left Heart | 707 | 2.3 | 271 | 78.6% | 15.1% | 6.3% |
| **Respiratory** | 2,187 | 7.2 | 1,405 | 52.2% | 39.6% | 8.2% |
| **Orofacial clefts** |  |  |  |  |  |  |
| Cleft Lip | 1,117 | 3.7 | 982 | 87.0% | 10.7% | 2.3% |
| Cleft Palate | 1,906 | 6.3 | 1,653 | 62.6% | 11.9% | 25.6% |
| Cleft Lip and Palate | 1,592 | 5.2 | 1,378 | 78.8% | 14.9% | 6.2% |
| **Digestive System** | 7,692 | 25.3 | 5,433 | 55.3% | 35.9% | 8.7% |
| Anorectal Malformations | 1,021 | 3.4 | 780 | 25.5% | 58.8% | 15.6% |
| Hirschsprung's Disease | 730 | 2.4 | 602 | 75.4% | 15.3% | 9.3% |
| Gastroschisis | 1,240 | 4.1 | 961 | 83.1% | ** | <1.0% |
| Unilateral Renal Agenesis | 583 | 1.9 | 453 | 71.3% | 22.5% | 6.2% |
| Congenital Hydronephrosis | 5,129 | 16.9 | 3,973 | 88.9% | 8.4% | 2.7% |
| Hypospadias | 5,118 | 16.8 | 4,329 | 76.3% | 9.6% | 14.2% |
| Club Foot - Talipes Equinovarus | 3,328 | 10.9 | 2,565 | 86.3% | 10.4% | 3.3% |
| Polydactyly | 5,129 | 16.9 | 4,027 | 86.4% | 6.4% | 7.2% |
| Syndactyly | 2,432 | 8.0 | 1,918 | 80.6% | 13.6% | 5.8% |
| Craniosynostosis | 934 | 3.1 | 766 | 68.7% | 16.4% | 14.9% |
| **Chromosomal Anomalies** |  |  |  |  |  |  |
| Down Syndrome | 3,023 | 9.9 | 2,134 | - | - | 100.0% |
| Turner Syndrome | 190 | 0.6 | 135 | - | - | 100.0% |
| Klinefelter Syndrome | 89 | 0.3 | 73 | - | - | 100.0% |
| Di George Syndrome | 198 | 0.7 | 136 | - | - | 100.0% |
| Karyotype XXX | 55 | 0.2 | 40 | - | - | 100.0% |

CA: congenital anomaly; CHD: congenital heart defect; HES: Hospital Episode Statistics; PDA: patent ductus arteriosus.
<sup>a</sup> as percentage of children included in study
\*\* not presented due to suppressed quantity in adjoining cell
- not applicable

Males comprised 51.4% of all children but 59.7% of those with CAs (Table 2). Compared to children without CAs, proportionally more children with CAs were born preterm (<37 weeks) and had low birthweight (<2500g). Children with CAs were more likely to have mothers who were <20 or ≥40 years, be of Asian/Chinese or Black ethnicity, have FSME, and be in the lowest two deprivation quintiles. Most variables had no or relatively little missing data (<4.0%), except for birthweight (23.0%) and gestational age (33.6%), with a slightly higher rate for children with CAs.

**Table 2:** Distribution of sociodemographic characteristics by congenital anomaly (CA) status

|  |  | No CA, n (%) | Any CA, n (%) | All, n (%) |
| --- | --- | --- | --- | --- |
| **TOTAL** |  | **2,272,742 (100.0)** | **78,847 (100.0)** | **2,351,589 (100.0)** |
| Year of birth | 2003/04 | 421,195 (18.5) | 14,483 (18.4) | 435,678 (18.5) |
|  | 2004/05 | 445,480 (19.6) | 15,311 (19.4) | 460,791 (19.6) |
|  | 2005/06 | 459,846 (20.2) | 15,920 (20.2) | 475,766 (20.2) |
|  | 2006/07 | 470,683 (20.7) | 16,283 (20.6) | 486,966 (20.7) |
|  | 2007/08 | 475,538 (20.9) | 16,850 (21.4) | 492,388 (20.9) |
| Sex at birth | Male | 1,160,586 (51.1) | 47,101 (59.7) | 1,207,687 (51.4) |
|  | Female | 1,112,156 (48.9) | 31,746 (40.3) | 1,143,902 (48.6) |
| Gestational Age, weeks | <32 | 10,687 (0.5) | 1,965 (2.5) | 12,652 (0.5) |
|  | 32-38 | 75,302 (3.3) | 4,802 (6.1) | 80,104 (3.4) |
|  | 37-41 | 1,355,335 (59.6) | 41,817 (53.0) | 1,397,152 (59.4) |
|  | 42+ | 69,391 (3.0) | 1,944 (2.5) | 71,335 (3.0) |
|  | Missing | 762,027 (33.5) | 28,319 (35.9) | 790,346 (33.6) |
| Birthweight, grams | <2500 | 96,228 (4.2) | 8,696 (11.0) | 104,924 (4.5) |
|  | 2500-3999 | 1,453,552 (64.0) | 44,758 (56.8) | 1,498,310 (63.7) |
|  | 4000+ | 201,694 (8.9) | 5,890 (7.5) | 207,584 (8.8) |
|  | Missing | 521,268 (22.9) | 19,503 (24.7) | 540,771 (23.0) |
| Maternal Age at birth, years | <20 | 155,599 (6.8) | 5,627 (7.1) | 161,226 (6.9) |
|  | 20-29 | 1,009,114 (44.4) | 34,321 (43.5) | 1,043,435 (44.4) |
|  | 30-34 | 610,174 (26.9) | 20,047 (25.4) | 630,221 (26.8) |
|  | 35-39 | 340,290 (15.0) | 11,739 (14.9) | 352,029 (15.0) |
|  | 40+ | 69,898 (3.1) | 2,902 (3.7) | 72,800 (3.1) |
|  | Missing | 87,667 (3.9) | 4,211 (5.3) | 91,878 (3.9) |
| Major Ethnic Group | White | 1,611,153 (70.9) | 54,834 (69.5) | 1,665,987 (70.8) |
|  | Black | 104,737 (4.6) | 3,833 (4.9) | 108,570 (4.6) |
|  | Asian/Chinese | 200,567 (8.8) | 7,765 (9.9) | 208,332 (8.9) |
|  | Mixed | 106,556 (4.7) | 3,668 (4.7) | 110,224 (4.7) |
|  | Other | 26,943 (1.2) | 853 (1.1) | 27,796 (1.2) |
|  | Unclassified | 222,786 (9.8) | 7,894 (10.0) | 230,680 (9.8) |
| Income Deprivation Affecting Children Index (IDACI) Quintile | Most Deprived | 596,343 (26.2) | 21,685 (27.5) | 618,028 (26.3) |
|  | 2nd most deprived | 473,994 (20.9) | 16,706 (21.2) | 490,700 (20.9) |
|  | Middle | 417,944 (18.4) | 14,267 (18.1) | 432,211 (18.4) |
|  | 2nd least deprived | 398,665 (17.5) | 13,195 (16.7) | 411,860 (17.5) |
|  | Least deprived | 378,084 (16.6) | 12,702 (16.1) | 390,786 (16.6) |
|  | Missing | 7,712 (0.3) | 292 (0.4) | 8,004 (0.3) |
| Free School Meals Eligibility (FSME) | No | 1,859,308 (81.8) | 63,082 (80.0) | 1,922,390 (81.8) |
|  | Yes | 413,434 (18.2) | 15,765 (20.0) | 429,199 (18.2) |
| Region | East Midlands | 197,047 (8.7) | 7,853 (10.0) | 204,900 (8.7) |
|  | East of England | 250,776 (11.0) | 7,096 (9.0) | 257,872 (11.0) |
|  | London | 349,531 (15.4) | 9,686 (12.3) | 359,217 (15.3) |
|  | North East | 117,115 (5.2) | 4,045 (5.1) | 121,160 (5.2) |
|  | North West | 295,778 (13.0) | 12,263 (15.6) | 308,041 (13.1) |
|  | South East | 357,730 (15.7) | 12,175 (15.4) | 369,905 (15.7) |
|  | South West | 218,526 (9.6) | 7,480 (9.5) | 226,006 (9.6) |
|  | West Midlands | 231,208 (10.2) | 10,675 (13.5) | 241,883 (10.3) |
|  | Yorkshire and The Humber | 247,535 (10.9) | 7,286 (9.2) | 254,821 (10.8) |
|  | Missing | 7,496 (0.3) | 288 (0.4) | 7,784 (0.3) |

*CA: congenital anomaly; FSME: Free School Meals Eligibility; IDACI: Income Deprivation Affecting Children Index*

### 3.1 | Good Level of Development

Overall, 57.0% of enrolled children without CAs reached GLD at EYFSP, compared with 45.7% of children with CAs (Figure 2). Children with isolated congenital hydronephrosis, club foot and polydactyly had the highest proportions reaching GLD (≥51.9%), slightly below their unaffected peers, whilst those with hypoplastic left heart and congenital hydrocephalus had the lowest. Excepting Karyotype XXX, syndromic CAs showed the lowest GLD achievement rates (<1% for Down syndrome). Sex differences existed across CA subgroups, with 39.8% of males reaching GLD compared with 54.5% of females on average (eTable4).

**Figure 2:**
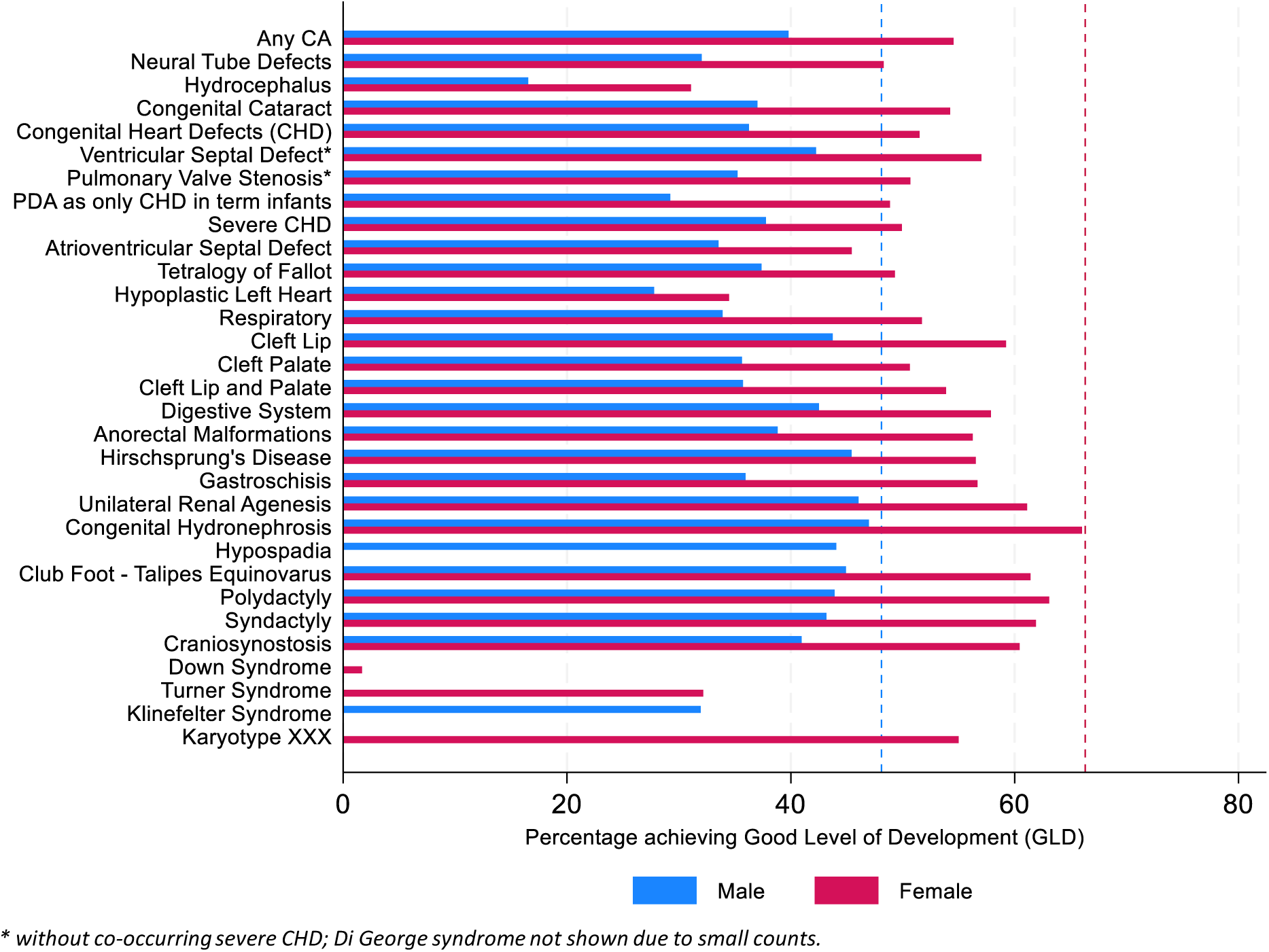
Percentage achieving Good Level of Development (GLD) at EYFSP, by sex and CA subgroup. Dashed lines represent values for children without CAs (blue=male; red=female)

### 3.2 | Subject attainment for English and Maths

The assessment rate for English and Maths for children without CAs decreased from 99.3% at EYFSP to 99.2% at KS1, and 97.2% at KS2, whereas for children with CAs it declined more sharply from 99.1% (EYFSP) to 98.6% (KS1) and 88.6% (KS2) (Table 3). Attainment rates for children with CAs in English rose from 56.9% (EYFSP) to 65.3% (KS2), which were consistently 11-12% lower than for their peers.

**Table 3:** Number of children enrolled, % assessed and % who reached expected level of attainment by Key Stage, subject and congenital anomaly (CA) subgroup.

| subgroup | EYFSP |  |  |  |  | KS1 |  |  |  |  | KS2 |  |  |  |  |
| --- | --- | --- | --- | --- | --- | --- | --- | --- | --- | --- | --- | --- | --- | --- | --- |
|  | N in census | English |  | Maths |  | N in census | English |  | Maths |  | N in census | English |  | Maths |  |
|  |  | % assessed | % expected | % assessed | % expected |  | % assessed | % expected | % assessed | % expected |  | % assessed | % expected | % assessed | % expected |
| All Children | 2,351,589 | 99.3 | 68.2 | 99.3 | 78.1 | 2,319,460 | 99.2 | 66.6 | 99.1 | 78.4 | 2,257,545 | 96.9 | 75.9 | 97.0 | 77.4 |
| No CA | 2,272,742 | 99.3 | 68.6 | 99.3 | 78.5 | 2,241,658 | 99.2 | 67.0 | 99.2 | 78.8 | 2,181,797 | 97.2 | 76.3 | 97.3 | 77.8 |
| Any CA | 78,847 | 99.1 | 56.9 | 99.1 | 67.6 | 77,802 | 98.9 | 55.4 | 98.6 | 66.8 | 75,748 | 88.7 | 65.3 | 88.6 | 66.4 |
| Neural Tube Defects | 275 | 98.5 | 50.2 | 98.5 | 63.3 | 272 | 99.6 | 54.0 | 98.9 | 59.9 | 266 | 85.0 | 59.8 | 84.6 | 57.9 |
| Hydrocephalus | 390 | 97.9 | 31.0 | 97.9 | 39.5 | 386 | 97.9 | 29.3 | 96.9 | 36.8 | 374 | 61.0 | 36.9 | 61.0 | 35.6 |
| Congenital Cataract | 382 | 99.0 | 58.9 | 99.0 | 70.9 | 377 | 99.5 | 57.6 | 99.5 | 69.2 | 363 | 89.3 | 67.5 | 90.4 | 67.2 |
| Congenital Heart Defects (CHD) | 11,368 | 99.2 | 54.2 | 99.2 | 65.3 | 11,222 | 98.8 | 52.3 | 98.6 | 63.3 | 10,925 | 89.8 | 62.9 | 89.7 | 62.6 |
| Ventricular Septal Defect* | 2,631 | 99.2 | 59.9 | 99.2 | 70.7 | 2,597 | 99.1 | 57.7 | 99.0 | 68.7 | 2,538 | 91.9 | 68.4 | 91.6 | 68.3 |
| Pulmonary Valve Stenosis* | 429 | 99.1 | 53.8 | 99.1 | 66.0 | 422 | 99.3 | 55.0 | 99.1 | 64.5 | 413 | 91.3 | 64.4 | 91.5 | 60.8 |
| PDA as only CHD in term infants | 2,068 | 98.9 | 49.4 | 98.9 | 60.7 | 2,047 | 98.5 | 47.8 | 98.3 | 57.4 | 2,001 | 88.6 | 61.1 | 87.9 | 56.5 |
| Severe CHD | 2,991 | 99.3 | 54.6 | 99.3 | 66.2 | 2,946 | 98.9 | 51.4 | 98.6 | 63.4 | 2,854 | 88.7 | 60.4 | 89.0 | 62.9 |
| Atrioventricular Septal Defect | 337 | 99.1 | 54.6 | 99.1 | 65.0 | 329 | 99.1 | 52.0 | 99.1 | 61.7 | 323 | 88.2 | 55.7 | 87.9 | 58.2 |
| Tetralogy of Fallot | 528 | 100.0 | 55.1 | 100.0 | 67.4 | 517 | 99.0 | 51.8 | 98.8 | 62.9 | 501 | 88.8 | 61.3 | 89.6 | 65.7 |
| Hypoplastic Left Heart | 213 | 98.1 | 41.3 | 98.1 | 56.3 | 208 | 98.1 | 39.9 | 98.1 | 51.0 | 196 | 82.7 | 51.5 | 80.6 | 47.4 |
| Respiratory | 734 | 98.8 | 52.2 | 98.8 | 63.6 | 723 | 98.6 | 52.3 | 98.6 | 63.2 | 706 | 89.0 | 62.2 | 89.2 | 63.2 |
| Cleft Lip | 854 | 99.1 | 61.4 | 99.1 | 73.5 | 837 | 99.5 | 61.6 | 99.3 | 72.8 | 817 | 94.5 | 70.0 | 95.3 | 75.3 |
| Cleft Palate | 1,034 | 99.8 | 54.6 | 99.8 | 66.1 | 1,023 | 99.6 | 53.3 | 99.4 | 64.5 | 1,001 | 91.4 | 65.3 | 92.5 | 65.4 |
| Cleft Lip and Palate | 1,086 | 99.4 | 54.3 | 99.4 | 66.9 | 1,075 | 99.3 | 52.3 | 99.2 | 67.3 | 1,048 | 93.0 | 64.6 | 94.1 | 66.6 |
| Digestive System | 3,007 | 99.3 | 60.2 | 99.3 | 72.0 | 2,977 | 99.5 | 59.4 | 99.4 | 72.1 | 2,901 | 93.5 | 69.8 | 93.4 | 71.3 |
| Anorectal Malformations | 199 | 99.0 | 57.8 | 99.0 | 68.8 | 196 | 99.5 | 59.2 | 99.5 | 69.9 | 192 | 91.7 | 70.8 | 93.2 | 69.3 |
| Hirschsprung's Disease | 454 | 99.6 | 59.7 | 99.6 | 73.6 | 448 | 99.6 | 58.0 | 99.3 | 72.8 | 437 | 93.1 | 69.6 | 92.7 | 71.9 |
| Gastroschisis | 799 | 99.1 | 57.6 | 99.1 | 68.1 | 795 | 99.2 | 54.2 | 99.1 | 66.7 | 784 | 96.0 | 65.6 | 96.2 | 67.0 |
| Unilateral Renal Agenesis | 323 | 98.1 | 61.6 | 98.1 | 73.7 | 316 | 98.4 | 66.5 | 98.4 | 77.2 | 312 | 93.3 | 66.3 | 93.6 | 72.8 |
| Congenital Hydronephrosis | 3,532 | 99.4 | 64.2 | 99.4 | 75.5 | 3,479 | 99.0 | 61.3 | 98.9 | 77.5 | 3,392 | 95.7 | 74.1 | 95.9 | 76.4 |
| Hypospadias | 3,302 | 99.2 | 57.8 | 99.2 | 71.8 | 3,265 | 99.2 | 55.5 | 99.2 | 73.8 | 3,195 | 94.3 | 69.5 | 94.5 | 74.2 |
| Club Foot - Talipes Equinovarus | 2,213 | 99.2 | 63.4 | 99.2 | 74.3 | 2,185 | 99.4 | 61.0 | 99.4 | 73.9 | 2,119 | 94.6 | 71.1 | 94.9 | 73.0 |
| Polydactyly | 3,480 | 99.2 | 63.0 | 99.2 | 74.3 | 3,433 | 99.1 | 63.2 | 99.1 | 74.2 | 3,345 | 95.0 | 72.5 | 95.0 | 74.1 |
| Syndactyly | 1,545 | 99.7 | 64.3 | 99.7 | 75.2 | 1,527 | 99.0 | 62.3 | 98.8 | 75.4 | 1,477 | 95.3 | 72.5 | 95.2 | 74.0 |
| Craniosynostosis | 526 | 99.2 | 60.5 | 99.2 | 73.0 | 516 | 99.0 | 57.6 | 98.6 | 69.4 | 502 | 91.6 | 69.1 | 91.4 | 68.5 |
| Down Syndrome | 2,134 | 97.4 | 2.7 | 97.4 | 4.0 | 2,113 | 95.1 | 0.7 | 91.7 | 0.7 | 2,083 | 3.2 | 0.9 | 2.9 | 0.7 |
| Turner Syndrome | 115 | 100.0 | 42.6 | 100.0 | 47.0 | 114 | 100.0 | 44.7 | 99.1 | 38.6 | 112 | 80.4 | 54.5 | 77.7 | 40.2 |
| Klinefelter Syndrome | 72 | 100.0 | 40.3 | 100.0 | 66.7 | 72 | 100.0 | 26.4 | 100.0 | 44.4 | 69 | 75.4 | 27.5 | 76.8 | 36.2 |
| Di George Syndrome | 136 | 98.5 | 8.8 | 98.5 | 10.3 | 135 | 97.0 | - | 96.3 | - | 132 | 39.4 | - | 37.1 | - |
| Karyotype XXX | 40 | 97.5 | 60.0 | 97.5 | 65.0 | 39 | 100.0 | 30.8 | 100.0 | 48.7 | 39 | 87.2 | 38.5 | 87.2 | 35.9 |

*CHD=congenital heart defect; GLD=Good Level of Development; EYFSP=Early Years Foundation Stage Profile; KS1=Key Stage 1; KS2=Key Stage 2; PDA=patent ductus arteriosus*
\* without co-occurring severe CHD
- suppressed due to small counts
% assessed = percentage of those in census who had an assessment
% expected = percentage of those in census who reached expected level of attainment

Attainment rates for Maths remained stable but a similar gap existed (66-67% for children with CAs, 77-78% for peers). Amongst children with isolated CAs, those with renal anomalies, limb defects and cleft lip had the highest attainment rates, whereas those with congenital hydrocephalus had the lowest. Children with patent ductus arteriosus (PDA), a non-severe heart defect, generally performed worse than children with severe CHD and many non-syndromic CAs at all ages. For syndromic CAs, children with Turner, Klinefelter and Karyotype XXX syndromes began with relatively high attainment rates, but the latter two declined at older ages. Under 1% of children with Down Syndrome achieved expected levels at KS1 and KS2.

Adjusting for sociodemographic factors, children with CAs were less likely than peers to achieve expected levels in English, (adjusted risk ratio, aRR (95%CI): 0.86 (0.85,0.86) [EYFSP and KS1]; 0.87 (0.87,0.88) [KS2]) (Table 4). Similar results were seen for Maths (Table 5). Males were less likely than females to achieve expected levels, with no variation across CA subgroups (aRR (95%CI): 0.81 (0.81,0.81) [EYFSP], 0.91 (0.90,0.91) [KS2] for English; 0.93 (0.92,0.93) [EYFSP], 0.99 (0.99,0.99) [KS2] for Maths).

**Table 4:** Adjusted risk ratios for reaching expected levels of attainment in English, comparing children with and without congenital anomalies (CAs), by key stage and CA subgroup. For each comparison, number of children without CAs=2,177,654

| subgroup | Cases, N | EYFSP |  | KS1 |  | KS2 |  |
| --- | --- | --- | --- | --- | --- | --- | --- |
|  |  | RR (95%CI) | RR (95%CI) | RR (95%CI) | RR (95%CI) | RR (95%CI) | RR (95%CI) |
|  |  | Sex adj. | Full adj. | Sex adj. | Full adj. | Sex adj. | Full adj. |
| Any CA | 74,357 | 0.85 (0.84,0.86) | 0.86 (0.85,0.86) | 0.85 (0.84,0.85) | 0.85 (0.85,0.86) | 0.87 (0.86,0.87) | 0.87 (0.87,0.88) |
| Neural Tube Defects | 248 | 0.73 (0.65,0.83) | 0.77 (0.68,0.86) | 0.80 (0.71,0.90) | 0.83 (0.74,0.94) | 0.81 (0.73,0.89) | 0.83 (0.75,0.92) |
| Hydrocephalus | 362 | 0.47 (0.41,0.55) | 0.49 (0.42,0.57) | 0.46 (0.40,0.54) | 0.48 (0.41,0.56) | 0.51 (0.44,0.58) | 0.52 (0.45,0.59) |
| Congenital Cataract | 362 | 0.85 (0.78,0.93) | 0.86 (0.79,0.93) | 0.86 (0.78,0.93) | 0.86 (0.79,0.94) | 0.87 (0.81,0.94) | 0.88 (0.81,0.94) |
| Congenital Heart Defects (CHD) | 10,216 | 0.81 (0.79,0.82) | 0.82 (0.80,0.83) | 0.80 (0.78,0.81) | 0.81 (0.79,0.82) | 0.83 (0.82,0.84) | 0.84 (0.83,0.85) |
| Ventricular Septal Defect | 2,498 | 0.88 (0.85,0.91) | 0.89 (0.86,0.92) | 0.86 (0.83,0.89) | 0.87 (0.84,0.90) | 0.91 (0.88,0.93) | 0.91 (0.89,0.94) |
| Pulmonary Valve Stenosis | 397 | 0.78 (0.71,0.85) | 0.78 (0.71,0.85) | 0.81 (0.74,0.89) | 0.82 (0.75,0.89) | 0.85 (0.79,0.92) | 0.86 (0.79,0.92) |
| PDA as only CHD in term infants | 1,610 | 0.74 (0.71,0.78) | 0.76 (0.72,0.79) | 0.74 (0.71,0.78) | 0.76 (0.72,0.80) | 0.82 (0.79,0.85) | 0.83 (0.80,0.86) |
| Severe CHD | 2,763 | 0.81 (0.79,0.84) | 0.82 (0.79,0.85) | 0.79 (0.77,0.82) | 0.80 (0.77,0.83) | 0.80 (0.77,0.82) | 0.80 (0.78,0.83) |
| Atrioventricular Septal Defect | 320 | 0.81 (0.73,0.89) | 0.82 (0.75,0.90) | 0.77 (0.70,0.86) | 0.79 (0.71,0.87) | 0.74 (0.67,0.82) | 0.75 (0.68,0.83) |
| Tetralogy of Fallot | 506 | 0.82 (0.76,0.89) | 0.83 (0.77,0.90) | 0.79 (0.73,0.86) | 0.79 (0.73,0.86) | 0.81 (0.75,0.87) | 0.81 (0.75,0.87) |
| Hypoplastic Left Heart | 194 | 0.62 (0.53,0.73) | 0.65 (0.55,0.76) | 0.62 (0.52,0.73) | 0.64 (0.54,0.76) | 0.66 (0.57,0.76) | 0.67 (0.58,0.78) |
| Respiratory | 665 | 0.78 (0.72,0.83) | 0.79 (0.73,0.84) | 0.79 (0.73,0.85) | 0.80 (0.74,0.86) | 0.82 (0.77,0.87) | 0.82 (0.77,0.87) |
| Cleft Lip | 825 | 0.92 (0.87,0.97) | 0.93 (0.88,0.98) | 0.95 (0.90,1.00) | 0.96 (0.91,1.01) | 0.93 (0.89,0.98) | 0.94 (0.90,0.99) |
| Cleft Palate | 989 | 0.79 (0.75,0.84) | 0.80 (0.76,0.84) | 0.79 (0.75,0.84) | 0.80 (0.76,0.85) | 0.87 (0.83,0.91) | 0.87 (0.83,0.91) |
| Cleft Lip and Palate | 1,044 | 0.81 (0.77,0.86) | 0.83 (0.78,0.87) | 0.81 (0.77,0.86) | 0.83 (0.78,0.88) | 0.86 (0.82,0.91) | 0.87 (0.83,0.92) |
| Digestive System | 2,748 | 0.88 (0.85,0.91) | 0.89 (0.86,0.91) | 0.90 (0.88,0.93) | 0.91 (0.88,0.94) | 0.92 (0.90,0.95) | 0.93 (0.90,0.95) |
| Anorectal Malformations | 172 | 0.85 (0.75,0.97) | 0.86 (0.76,0.97) | 0.94 (0.83,1.05) | 0.94 (0.84,1.06) | 0.95 (0.86,1.05) | 0.95 (0.86,1.05) |
| Hirschsprung's Disease | 411 | 0.93 (0.86,1.00) | 0.94 (0.87,1.01) | 0.92 (0.85,1.00) | 0.93 (0.86,1.01) | 0.94 (0.87,1.00) | 0.94 (0.88,1.01) |
| Gastroschisis | 705 | 0.83 (0.78,0.88) | 0.92 (0.87,0.98) | 0.81 (0.76,0.87) | 0.92 (0.86,0.99) | 0.87 (0.82,0.92) | 0.94 (0.89,0.99) |
| Unilateral Renal Agenesis | 304 | 0.93 (0.85,1.02) | 0.94 (0.86,1.02) | 1.03 (0.95,1.11) | 1.04 (0.96,1.12) | 0.89 (0.82,0.97) | 0.90 (0.82,0.97) |
| Congenital Hydronephrosis | 3,405 | 0.98 (0.96,1.01) | 0.97 (0.95,1.00) | 0.97 (0.94,0.99) | 0.96 (0.93,0.98) | 1.00 (0.98,1.02) | 0.99 (0.97,1.01) |
| Hypospadias | 3,170 | 0.94 (0.91,0.97) | 0.94 (0.92,0.97) | 0.94 (0.91,0.97) | 0.94 (0.91,0.97) | 0.97 (0.94,0.99) | 0.97 (0.95,0.99) |
| Club Foot - Talipes Equinovarus | 2,145 | 0.94 (0.91,0.97) | 0.94 (0.91,0.97) | 0.93 (0.90,0.96) | 0.93 (0.90,0.97) | 0.94 (0.91,0.97) | 0.94 (0.91,0.97) |
| Polydactyly | 3,352 | 0.93 (0.91,0.95) | 0.96 (0.94,0.99) | 0.96 (0.93,0.98) | 0.97 (0.95,1.00) | 0.96 (0.94,0.98) | 0.97 (0.95,0.99) |
| Syndactyly | 1,504 | 0.97 (0.93,1.00) | 0.95 (0.91,0.98) | 0.97 (0.93,1.01) | 0.96 (0.92,0.99) | 0.96 (0.93,1.00) | 0.96 (0.93,0.99) |
| Craniosynostosis | 502 | 0.90 (0.84,0.97) | 0.89 (0.83,0.95) | 0.87 (0.81,0.94) | 0.86 (0.80,0.93) | 0.91 (0.86,0.97) | 0.90 (0.85,0.96) |
| Down Syndrome | 1,992 | 0.04 (0.03,0.05) | 0.04 (0.03,0.05) | 0.01 (0.01,0.02) | 0.01 (0.01,0.02) | 0.02 (0.01,0.03) | 0.02 (0.01,0.03) |
| Turner Syndrome | 125 | 0.52 (0.42,0.65) | 0.52 (0.42,0.64) | 0.55 (0.44,0.68) | 0.55 (0.44,0.68) | 0.63 (0.53,0.76) | 0.63 (0.53,0.76) |
| Klinefelter Syndrome | 69 | 0.66 (0.49,0.87) | 0.63 (0.47,0.83) | 0.42 (0.28,0.63) | 0.40 (0.27,0.61) | 0.37 (0.25,0.56) | 0.36 (0.24,0.54) |
| Di George Syndrome | 126 | 0.15 (0.09,0.25) | 0.15 (0.09,0.26) | 0.10 (0.05,0.19) | 0.10 (0.05,0.19) | 0.12 (0.07,0.21) | 0.12 (0.07,0.21) |
| Karyotype XXX | 36 | 0.81 (0.62,1.05) | 0.74 (0.57,0.96) | 0.38 (0.22,0.64) | 0.35 (0.21,0.59) | 0.47 (0.30,0.72) | 0.44 (0.29,0.69) |

*CA=congenital anomaly; CI=confidence interval; EYFSP=Early Years Foundation Stage Profile; KS1=Key Stage 1; KS2=Key Stage 2; PDA=patent ductus arteriosus; RR=risk ratio; Sex adj.= adjusted for sex at birth; Full adj.=adjusted for sex, maternal age at birth, ethnicity, income deprivation affecting children index (IDACI) quintile, free school meals eligibility (FSME)*

**Table 5:** Adjusted risk ratios for reaching expected levels of attainment in Maths, comparing children with and without congenital anomalies (CAs), by key stage and CA subgroup. For each comparison, number of children without CAs=2,177,654

| subgroup | Cases, N | EYFSP |  | KS1 |  | KS2 |  |
| --- | --- | --- | --- | --- | --- | --- | --- |
|  |  | RR (95%CI) | RR (95%CI) | RR (95%CI) | RR (95%CI) | RR (95%CI) | RR (95%CI) |
|  |  | Sex adj. | Full adj. | Sex adj. | Full adj. | Sex adj. | Full adj. |
| Any CA | 74,357 | 0.87 (0.87,0.88) | 0.88 (0.87,0.88) | 0.86 (0.85,0.86) | 0.86 (0.86,0.86) | 0.86 (0.85,0.86) | 0.86 (0.86,0.87) |
| Neural Tube Defects | 248 | 0.81 (0.74,0.89) | 0.84 (0.76,0.92) | 0.78 (0.70,0.86) | 0.80 (0.72,0.88) | 0.76 (0.68,0.85) | 0.78 (0.70,0.86) |
| Hydrocephalus | 362 | 0.52 (0.46,0.59) | 0.53 (0.47,0.60) | 0.49 (0.43,0.55) | 0.50 (0.44,0.57) | 0.47 (0.41,0.54) | 0.48 (0.42,0.55) |
| Congenital Cataract | 362 | 0.89 (0.84,0.96) | 0.90 (0.85,0.96) | 0.88 (0.82,0.94) | 0.88 (0.82,0.94) | 0.85 (0.79,0.92) | 0.85 (0.79,0.92) |
| Congenital Heart Defects (CHD) | 10,216 | 0.85 (0.83,0.86) | 0.86 (0.84,0.87) | 0.81 (0.80,0.82) | 0.82 (0.81,0.83) | 0.82 (0.80,0.83) | 0.82 (0.81,0.84) |
| Ventricular Septal Defect | 2,498 | 0.91 (0.88,0.93) | 0.91 (0.89,0.93) | 0.88 (0.85,0.90) | 0.88 (0.86,0.91) | 0.89 (0.86,0.91) | 0.89 (0.87,0.92) |
| Pulmonary Valve Stenosis | 397 | 0.84 (0.78,0.90) | 0.84 (0.79,0.90) | 0.81 (0.75,0.87) | 0.81 (0.76,0.88) | 0.77 (0.71,0.84) | 0.78 (0.71,0.84) |
| PDA as only CHD in term infants | 1,610 | 0.79 (0.76,0.82) | 0.81 (0.78,0.84) | 0.76 (0.73,0.79) | 0.77 (0.74,0.80) | 0.76 (0.73,0.79) | 0.77 (0.73,0.80) |
| Severe CHD | 2,763 | 0.86 (0.83,0.88) | 0.86 (0.84,0.88) | 0.81 (0.78,0.83) | 0.81 (0.79,0.83) | 0.81 (0.79,0.83) | 0.81 (0.79,0.84) |
| Atrioventricular Septal Defect | 320 | 0.83 (0.77,0.90) | 0.85 (0.78,0.91) | 0.79 (0.72,0.86) | 0.80 (0.73,0.87) | 0.76 (0.69,0.84) | 0.77 (0.70,0.84) |
| Tetralogy of Fallot | 506 | 0.87 (0.82,0.92) | 0.88 (0.83,0.93) | 0.80 (0.75,0.85) | 0.80 (0.75,0.86) | 0.84 (0.79,0.90) | 0.84 (0.79,0.90) |
| Hypoplastic Left Heart | 194 | 0.72 (0.64,0.81) | 0.74 (0.65,0.83) | 0.66 (0.57,0.75) | 0.67 (0.59,0.77) | 0.61 (0.52,0.71) | 0.62 (0.53,0.72) |
| Respiratory | 665 | 0.81 (0.77,0.86) | 0.82 (0.78,0.87) | 0.80 (0.75,0.85) | 0.81 (0.76,0.86) | 0.81 (0.76,0.86) | 0.81 (0.76,0.86) |
| Cleft Lip | 825 | 0.95 (0.91,0.99) | 0.95 (0.91,0.99) | 0.92 (0.88,0.96) | 0.93 (0.89,0.97) | 0.97 (0.93,1.01) | 0.98 (0.94,1.02) |
| Cleft Palate | 989 | 0.84 (0.81,0.88) | 0.85 (0.81,0.88) | 0.82 (0.78,0.86) | 0.82 (0.79,0.86) | 0.86 (0.82,0.90) | 0.86 (0.82,0.90) |
| Cleft Lip and Palate | 1,044 | 0.86 (0.83,0.90) | 0.87 (0.84,0.91) | 0.87 (0.83,0.91) | 0.88 (0.84,0.91) | 0.87 (0.83,0.91) | 0.88 (0.84,0.92) |
| Digestive System | 2,748 | 0.92 (0.90,0.94) | 0.93 (0.91,0.95) | 0.92 (0.90,0.94) | 0.93 (0.91,0.95) | 0.92 (0.90,0.95) | 0.93 (0.90,0.95) |
| Anorectal Malformations | 172 | 0.90 (0.82,0.99) | 0.90 (0.82,0.99) | 0.90 (0.82,1.00) | 0.90 (0.82,1.00) | 0.91 (0.82,1.01) | 0.91 (0.82,1.01) |
| Hirschsprung's Disease | 411 | 0.95 (0.90,1.01) | 0.96 (0.91,1.02) | 0.95 (0.89,1.00) | 0.95 (0.90,1.01) | 0.93 (0.87,0.99) | 0.94 (0.88,1.00) |
| Gastroschisis | 705 | 0.87 (0.83,0.92) | 0.94 (0.90,0.99) | 0.85 (0.80,0.89) | 0.91 (0.87,0.96) | 0.87 (0.82,0.92) | 0.94 (0.89,0.99) |
| Unilateral Renal Agenesis | 304 | 0.95 (0.89,1.02) | 0.96 (0.89,1.02) | 0.98 (0.92,1.04) | 0.98 (0.92,1.05) | 0.94 (0.88,1.01) | 0.95 (0.88,1.02) |
| Congenital Hydronephrosis | 3,405 | 0.98 (0.96,1.00) | 0.97 (0.96,0.99) | 0.99 (0.97,1.01) | 0.99 (0.97,1.01) | 0.99 (0.97,1.01) | 0.98 (0.96,1.00) |
| Hypospadias | 3,170 | 0.95 (0.93,0.97) | 0.95 (0.93,0.97) | 0.96 (0.94,0.98) | 0.96 (0.94,0.98) | 0.97 (0.95,0.99) | 0.97 (0.95,0.99) |
| Club Foot - Talipes Equinovarus | 2,145 | 0.95 (0.93,0.98) | 0.95 (0.93,0.97) | 0.95 (0.92,0.97) | 0.95 (0.93,0.97) | 0.94 (0.91,0.96) | 0.94 (0.91,0.97) |
| Polydactyly | 3,352 | 0.95 (0.93,0.97) | 0.98 (0.96,1.00) | 0.94 (0.93,0.96) | 0.97 (0.95,0.99) | 0.95 (0.93,0.97) | 0.96 (0.94,0.98) |
| Syndactyly | 1,504 | 0.97 (0.94,1.00) | 0.95 (0.93,0.98) | 0.97 (0.94,1.00) | 0.96 (0.93,0.98) | 0.95 (0.92,0.98) | 0.95 (0.92,0.98) |
| Craniosynostosis | 502 | 0.94 (0.89,0.99) | 0.92 (0.88,0.97) | 0.87 (0.82,0.93) | 0.86 (0.82,0.92) | 0.86 (0.81,0.92) | 0.86 (0.81,0.92) |
| Down Syndrome | 1,992 | 0.05 (0.04,0.06) | 0.05 (0.04,0.06) | 0.01 (0.01,0.02) | 0.01 (0.01,0.02) | 0.01 (0.01,0.02) | 0.01 (0.01,0.02) |
| Turner Syndrome | 125 | 0.57 (0.47,0.68) | 0.56 (0.47,0.68) | 0.46 (0.36,0.58) | 0.46 (0.36,0.58) | 0.47 (0.37,0.59) | 0.47 (0.37,0.60) |
| Klinefelter Syndrome | 69 | 0.88 (0.75,1.04) | 0.85 (0.72,1.00) | 0.55 (0.42,0.73) | 0.54 (0.41,0.71) | 0.47 (0.34,0.65) | 0.46 (0.34,0.64) |
| Di George Syndrome | 126 | 0.17 (0.11,0.27) | 0.17 (0.11,0.27) | 0.11 (0.06,0.20) | 0.11 (0.06,0.20) | 0.11 (0.06,0.19) | 0.11 (0.06,0.19) |
| Karyotype XXX | 36 | 0.82 (0.65,1.03) | 0.77 (0.61,0.95) | 0.59 (0.42,0.84) | 0.57 (0.40,0.80) | 0.44 (0.28,0.70) | 0.43 (0.27,0.68) |

*CA=congenital anomaly; CI=confidence interval; EYFSP=Early Years Foundation Stage Profile; KS1=Key Stage 1; KS2=Key Stage 2; PDA=patent ductus arteriosus; RR=risk ratio; Sex adj.= adjusted for sex at birth; Full adj.=adjusted for sex, maternal age at birth, ethnicity, income deprivation affecting children index (IDACI) quintile, free school meals eligibility (FSME)*

The attainment rates for specific subgroups defined by EUROCAT and alternative codelists differed by ≤1%, with some exceptions in severe CHD (eTable5). Amongst children with CAs, attainment rates were highest for isolated, followed by multiple then genetic CAs across subjects and ages (eTable6). For the subset of children assessed at all ages, adjusted risk ratios for achieving expected levels, comparing children with and without CAs, were closer to the null (eTable7, eTable8).

### 3.3 | Standardised scores (z-scores) for English and Maths

On average, z-scores were lower by 0.36 (EYFSP), 0.39 (KS1) and 0.16 (KS2) points and showed greater variation for children with CAs, compared with peers (eTable9); a unit z-score represented approximately 4-6 scale points at EYFSP, 4 points at KS1 and 9 marks at KS2. Differences with peers were smallest for children with renal anomalies, cleft lip and polydactyly. Excepting Klinefelter and Karyotype XXX syndromes, mean scores for children with CAs peaked at KS2, although KS2 samples were smaller, particularly for Down Syndrome (3% of EYFSP sample).

Estimated mean z-scores decreased for females and stayed constant for males over time, with adjustment for sociodemographic factors having little impact (Table 6). Overall, the gap between females with and without CAs widened from -0.35 at EYFSP to -0.38 at KS2 for Maths (−0.32 and -0.31 respectively for English). For males, the difference between children with CAs and peers very slightly reduced for English and increased for Maths at KS2. Most of the variation was at age 5 rather than in subsequent rates of change (variance_intercept_=0.64 [95%CI: 0.64,0.64]; variance_slopes_=0.08 [95%CI: 0.08,0.08]). Figure 3 shows the predicted trajectories stratified by CA (any) and sex, and Figure 4 shows trajectories for selected CAs (sexes combined).

**Table 6:** Mean differences in standardised English and Maths scores by sex and congenital anomaly (CA) status estimated by linear mixed models using all children with CAs and 25% sample of children without CAs.

|  |  | EYFSP | KS1 | KS2 |
| --- | --- | --- | --- | --- |
| *Unadjusted* |  |  |  |  |
| Female | No CA | 0 | 0.01 (0.00, 0.01) | -0.10 (-0.11, -0.10) |
|  | Any CA | -0.33 (-0.34, -0.32) | -0.33 (-0.34, -0.32) | -0.43 (-0.44, -0.42) |
| Male | No CA | -0.34 (-0.34, -0.33) | -0.32 (-0.32, -0.31) | -0.34 (-0.35, -0.34) |
|  | Any CA | -0.67 (-0.68, -0.66) | -0.65 (-0.66, -0.65) | -0.67 (-0.68, -0.66) |
| *Adjusted for maternal age, ethnicity, IDACI quintile and FSME* |  |  |  |  |
| Female | No CA | 0 | 0.01 (0.00, 0.01) | -0.10 (-0.11, -0.10) |
|  | Any CA | -0.32 (-0.33, -0.31) | -0.32 (-0.33, -0.31) | -0.41 (-0.42, -0.40) |
| Male | No CA | -0.34 (-0.34, -0.33) | -0.32 (-0.32, -0.31) | -0.34 (-0.35, -0.34) |
|  | Any CA | -0.66 (-0.66, -0.65) | -0.64 (-0.65, -0.63) | -0.65 (-0.66, -0.64) |

**Maths** (total individuals 617,463)
|  |  | EYFSP | KS1 | KS2 |
| --- | --- | --- | --- | --- |
| *Unadjusted* |  |  |  |  |
| Female | No CA | 0 | -0.05 (-0.06, -0.05) | -0.15 (-0.15, -0.15) |
|  | Any CA | -0.35 (-0.36, -0.34) | -0.44 (-0.44, -0.43) | -0.53 (-0.54, -0.52) |
| Male | No CA | -0.12 (-0.13, -0.12) | -0.06 (-0.07, -0.06) | -0.10 (-0.11, -0.10) |
|  | Any CA | -0.47 (-0.48, -0.46) | -0.44 (-0.45, -0.43) | -0.48 (-0.49, -0.47) |
| *Adjusted for maternal age, ethnicity, IDACI quintile and FSME* |  |  |  |  |
| Female | No CA | 0 | -0.05 (-0.06, -0.05) | -0.15 (-0.15, -0.15) |
|  | Any CA | -0.34 (-0.35, -0.33) | -0.42 (-0.43, -0.41) | -0.51 (-0.52, -0.50) |
| Male | No CA | -0.12 (-0.13, -0.12) | -0.06 (-0.07, -0.06) | -0.11 (-0.11, -0.10) |
|  | Any CA | -0.46 (-0.47, -0.45) | -0.43 (-0.44, -0.42) | -0.47 (-0.48, -0.46) |

CA = congenital anomaly; EYFSP=Early Years Foundation Stage Profile; FSME=Free school meals eligibility; IDACI=Income deprivation affecting children index; KS1=Key Stage 1; KS2=Key Stage 2

**Figure 3:**
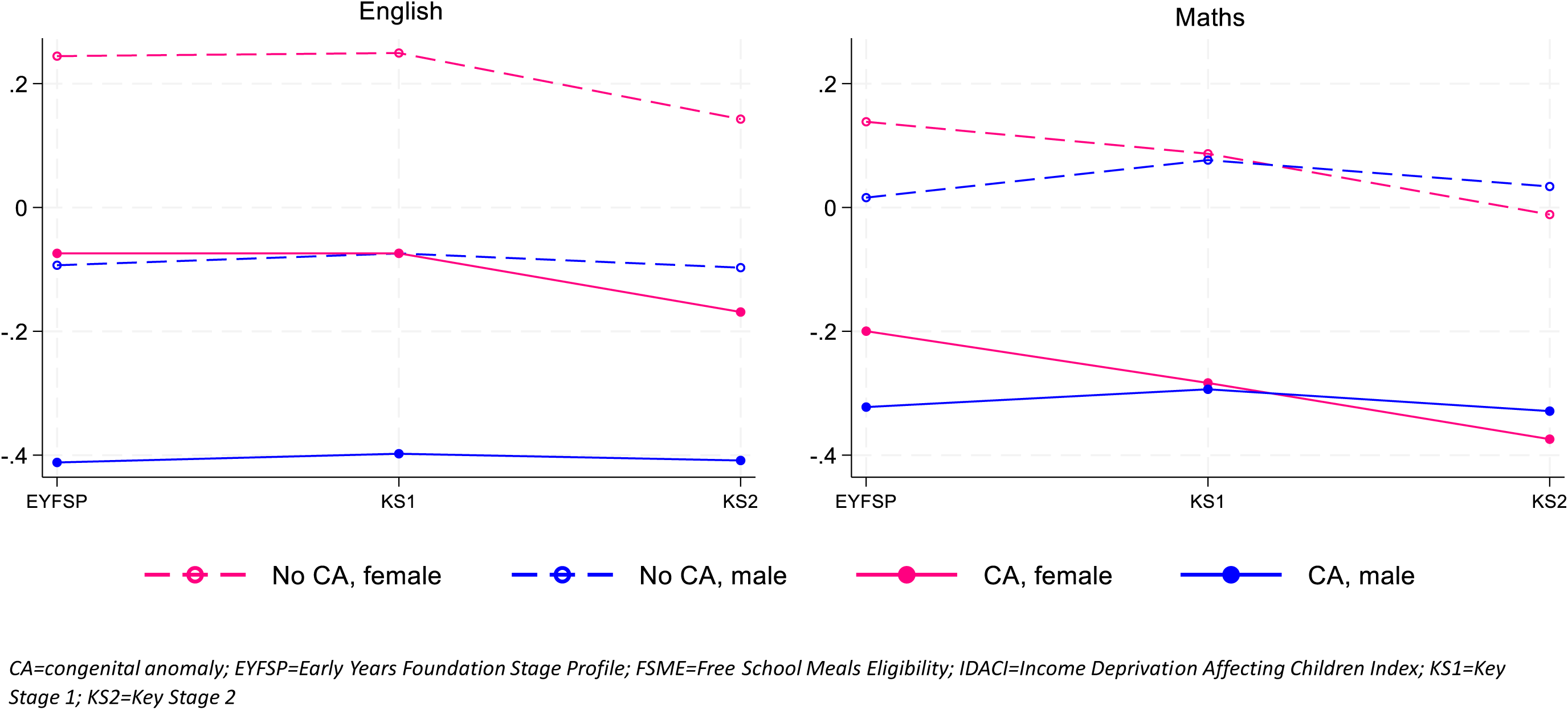
Estimated trajectories of English and Maths mean z-scores by categories of CA status and sex using linear mixed effects regression. Plots constructed using modal values of other adjusted covariates (maternal age: 20-29 years; ethnicity: White; IDACI quintile: 3rd quintile (middle); FSME: No)

**Figure 4:**
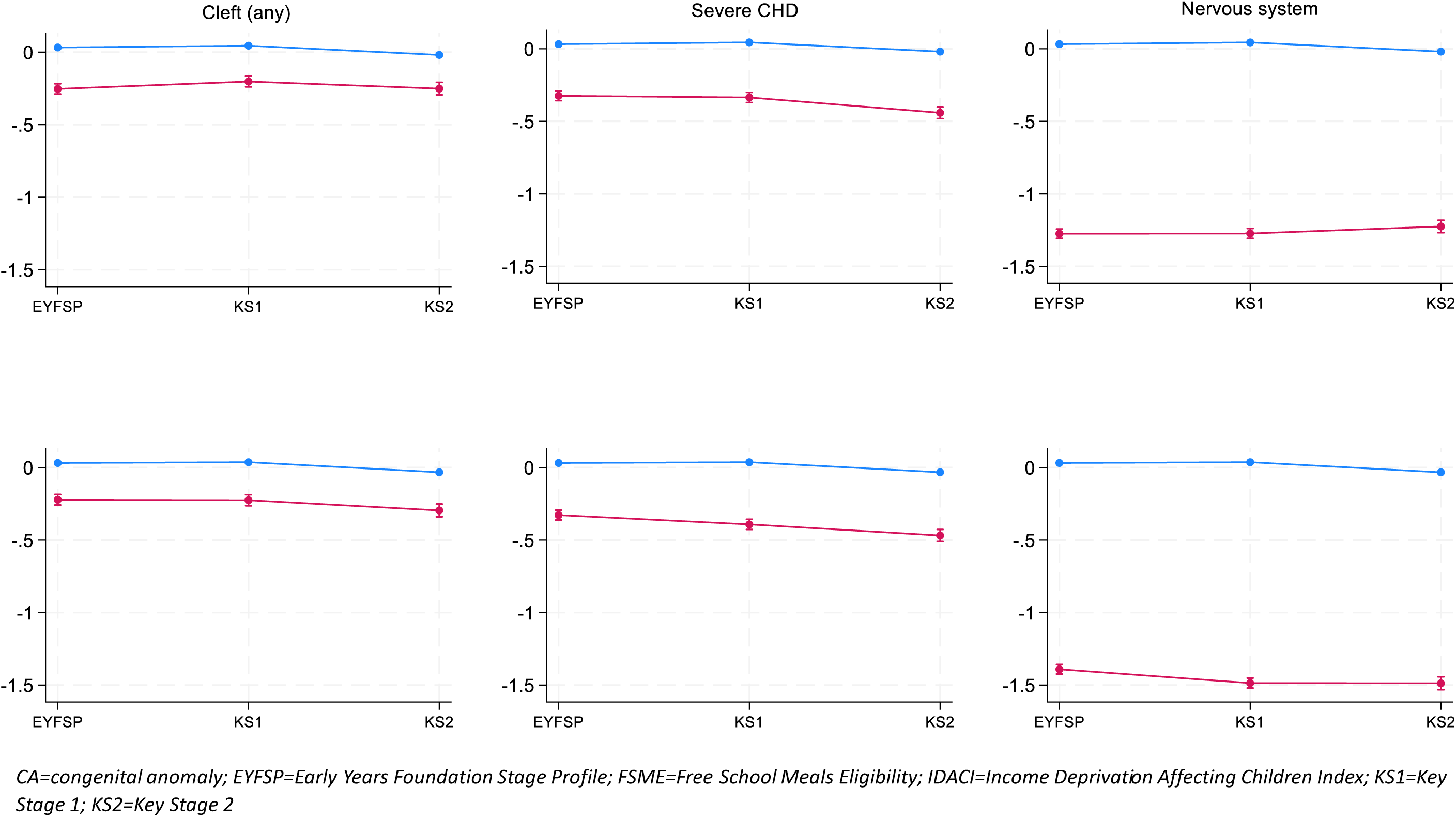
Estimated trajectories of mean standardised scores and 95% confidence intervals for English (top row) and Maths (bottom row), comparing children without CA (blue) and those with selected CAs (red). Plotted using average values of adjusted covariates (sex, maternal

## 4 | COMMENT

### 4.1 | Principal Findings

Amongst children with CAs enrolled at age 5, 45% reached GLD overall, with variations by CA subgroup and sex. This was 11% fewer than children without CAs, similar to the observed gaps for achieving expected levels in English and Maths at all ages. Proportionally more children with CAs were not assessed at age 11 compared with their peers (11% versus 3%). Adjusting for sociodemographic factors, children with CAs were on average 12-15% less likely than peers to reach expected attainment. Attainment for children with cleft lip, renal and limb anomalies were generally comparable with peers.

### 4.2 | Strengths of the study

HES covers 96.4% of births in England and provided a large sample size for our study.^24^ Up to 99% of NHS-funded hospital admissions were captured, enabling a range of CAs to be studied and compared with significant precision. Linkage to the NPD, containing information on pupils in state-funded schools (∼93% of total),^13^ provided insights into the academic journey and potential of children with CAs. Hence our findings are representative of a substantial part of the school-age population in England.

### 4.3 | Limitations of the Data

About 14% of total births could not be linked to the NPD, primarily due to the NPD not containing individual-level data on home-schooled children or those attending independent schools. Choices are shaped by parents’ preferences and circumstances, although admittedly some may be due to special educational needs that cannot be adequately met in state-funded settings. Additional factors include a combination of early exits (births to temporarily-resident mothers or emigration), linkage errors or incomplete matching identifiers.^25^ Given the diverse factors and comparatively small proportion, we expect the net influence on our results to be modest.

Misclassification of CA status is a limitation. In contrast to CA surveillance registries,^26^ where cases are notified by care teams and clinically reviewed, case ascertainment using administrative data relies on applying phenotype codelists deterministically to diagnosis and procedure codes, recorded primarily for reimbursement of healthcare provided. Children with CAs not requiring inpatient care could be missed (false negatives), whilst recording of suspected/differential diagnosis codes could generate false positives. Our estimated CA prevalence was 1.5% higher than EUROCAT statistics, and although CA registries could under-ascertain CAs, this discrepancy should be examined in future work. Where available, we have used alternative codelists which yielded more conservative prevalences to minimise the false positive rate. This may bias results toward more severe cases, but could help ascertain the upper limit of group differences; for selected CAs, attainment rates by alternative methods seemed very similar.

### 4.4 | Interpretation

Our aim was to provide evidence on the prognostic attainment trajectories in children with different CAs over the first seven years of their educational journey. Attainment rates peaked at KS2 for English and stayed largely constant for Maths, but the gap between children with CAs versus peers remained remarkably consistent at 11% lower throughout. The relatively sharp decrease in proportion of children with CAs assessed at KS2 is partially explained by transfers to special schools or disapplication from the National Curriculum after KS1. After controlling for sociodemographic factors, the association between CAs and risk of lower attainment was mostly unchanged across key stages. This was also true of children with CAs who sat all assessments, albeit differences with their peers were more attenuated.

An earlier cohort (1994-2004) from regional English CA registries linked to the NPD found that 72% and 73% of children with an isolated CA achieved expected levels in English and Maths at KS2 respectively, 6-7% fewer compared to peers.^9^ Our wider gaps of 11-12% are chiefly attributable to our overall CA group comprising not only isolated but also potential multiple and syndromic CAs, well-established to be associated with lower academic achievement,^4^ and secondarily to the different denominators used (number of children assessed versus enrolled). Once these have been accounted for, our findings are mutually reinforcing. This study found that the association of sex with attainment in children with CAs broadly mirrored that of children without CAs, with males performing worse at earlier stages but narrowing the gap with females by KS2 across all CA subgroups, particularly in English.

Our results also align with those of Park et al. on subject z-scores in children reported to the Cleft Registry and Audit Network.^3^ Children with cleft lip had the highest scores, followed by those with cleft lip and palate, then cleft palate, a pattern which we observed as well. They estimated that children with isolated clefts scored up to -0.29 (95% CI: -0.36, -0.22) lower than the national average in English, Maths and Science across all ages. Another study of CA cases from a single hospital found that 56-59% and 62% of children with CAs achieved expected attainment in English and Maths respectively at KS1.^4^ Allowing for differences in study settings and coding of outcome metrics, our findings were largely compatible, which gives assurance regarding the reliability and reproducibility of our results.

A pooled analysis using linked data from CA registries in Europe showed that children with CAs were at higher risk of co-morbidities such as cerebral palsy, seizures, hearing loss and visual impairment between ages 0-9.^27^ For example, the prevalence of visual impairment and cerebral palsy was on average 30 and 15 times higher respectively in children with CAs than those without CAs; nervous system CAs such as hydrocephalus were the most susceptible, but even isolated CHD, cleft palate and congenital hydronephrosis showed elevated rates of seizures, epilepsy and cerebral palsy. This may partly explain the unexpected lower attainment seen with some CA subgroups, e.g. PDA.

The current classification of CA subgroups does not fully capture variations in disease severity, in part due to the lack of granularity in the data. Additional factors include unidentified, or unidentifiable given available data, underlying learning or cognitive deficits. A greater proportion of children with Cas receive Special Educational Needs provision compared with their peers,^17, 28^ but this lies beyond our current scope and is being investigated elsewhere within the HOPE programme.^29^ Yet non-clinical drivers for lower academic achievement should not be overlooked. These may include unconscious bias or presuppositions about the academic potential of children with CA, which could predispose to self-fulfilling outcomes. Residual confounding by parental and familial factors, the home environment, as well as wider drivers of social disadvantage, are also important to explore in further analyses of ECHILD data.

### 4.5 | Conclusions

Of those enrolled at age 5 in state-funded schools in England, the proportions who continue to age 11 were similar between children with and without CAs. Children with CAs were however less likely to have curriculum assessments, or to reach expected levels of attainment, at every key stage. Notwithstanding some children with CAs who performed relatively well throughout, a significant minority did not participate in assessments after age 7. This highlights the need for close monitoring of children in this group, and for additional support to be provided, starting from an early age. With appropriate intervention, more children can be supported to advance the requisite skills and knowledge for secondary school. Some, but not all, of these advances will be measurable through curriculum assessments, and Government should consider how to best evaluate the benefits of early support for children’s development.

## Acknowledgements

We are grateful to all children and families whose de-identified data are used in this research. We thank the Patient and Public representative groups who have helped to develop this study, and the wider HOPE study and ECHILD management teams for their support for this work. We acknowledge the previous work by others who had developed methods for coding congenital anomalies cited in this study, as well as the EUROCAT network and EUROlinkCAT consortium for sharing their expertise and advice. Funding from the NIHR HOPE (Health Outcomes of young People in Education) study, the NIHR Great Ormond Street Hospital Biomedical Research Centre, and Health Data Research UK (grant number LOND1), contributed to this work. ECHILD is supported by Administrative Data Research UK and the Economic and Social Research Council (part of UK Research and Innovation) (grant numbers ES/V000977/1, ES/X003663/1, ES/X000427/1).

The education data analysed for this publication have been extracted from the National Pupil Database (NPD) which is compiled and owned by the Department for Education (DfE). DfE does not accept responsibility for any inferences or conclusions derived from the DfE Data Extracts by third parties.

This work contains statistical data from ONS which is Crown Copyright. The use of the ONS statistical data in this work does not imply the endorsement of the ONS in relation to the interpretation or analysis of the statistical data. This work uses research datasets which may not exactly reproduce National Statistics aggregates. The analysis was carried out in the Secure Research Service, part of the Office for National Statistics.

## Data Availability

ECHILD data are being made available to accredited researchers for research that benefits the provision of healthcare and education in England. Permission to access the ECHILD database is via application to the ECHILD team. Data can only be processed within the Office for National Statistics Secure Research Service by researchers who have undergone training through the Research Accreditation Service.

