## Supplemental Tables for "Educational attainment of children with major congenital anomalies during primary school in England: a population cohort study using linked administrative data from ECHILD"

### Supplementary Material

#### eTable1: Description of congenital anomaly (CA) subgroups, associated ICD-10 codes, exclusions, and published identification methods developed for Hospital Episode Statistics (HES)

| CA subgroups[c] | ICD-10 codes[d] (EUROCAT Guide 1.4) | EUROCAT exclusions | Published methods for HES data (alternative codelists) |
|---|---|---|---|
| **Any CA[a]** | Q-chapter, D215, D821, P350, P351, P371 | Exclude all minor anomalies as specified in Guide 1.4 | |
| **Neural Tube Defects (NTD)** | Q00, Q01, Q05 | | |
| Congenital hydrocephalus | Q03 | Exclude hydranencephaly. Exclude association with NTD subgroups | |
| Congenital cataract | Q120 | | |
| **Congenital Heart Defects (CHD)** | | | |
| **All CHD** | Q20-Q26 | Exclude patent ductus arteriosus (PDA) (Q250) with gestational age (GA) <37 weeks Exclude peripheral pulmonary artery stenosis (Q256) with GA <37 weeks | |
| Ventricular septal defect without severe CHD | Q210 | Exclude if severe CHD present | |
| Pulmonary valve stenosis without severe CHD | Q221 | Exclude if severe CHD present | |
| Patent ductus arteriosus (PDA) as the only CHD in term livebirths (GA 37+ weeks) | Q250 | Exclude if GA<37 weeks. Exclude if another CHD present | |
| **Severe CHD** | Q200, Q201, Q203, Q204, Q212, Q213, Q220, Q224, Q225, Q226, Q230, Q232, Q233, Q234, Q251, Q252, Q262 | | As EUROCAT and additional procedure codes as described in Gimeno et al., 2023[1] |
| Atrioventricular septal defect[b] | Q212 | | |
| Tetralogy of Fallot[b] | Q213 | | |
| Hypoplastic left heart[b] | Q234 | | |
| **Oro-facial clefts** | Q35-Q37 | Exclude if associated with holoprosencephaly or anencephaly, or Q357 | Diagnosis code + procedure code for primary cleft repair (F031 or F291); see Fitzsimons et al., 2018[2] |
| Cleft lip | Q36 | Exclude if associated with | As above |

| | | | |
|---|---|---|---|
| | | holoprosencephaly or anencephaly | |
| Cleft palate | Q35 | Exclude if associated with holoprosencephaly or anencephaly, or Q357 | As above |
| Cleft lip with cleft palate | Q37 | Exclude if associated with holoprosencephaly or anencephaly, or Q357 | As above |
| **Respiratory** | Q300, Q32-Q34 | Exclude Q336 | |
| **Digestive System** | Q38-Q45, Q790 | Exclude Q381, Q382, Q400, Q401, Q430, Q444 | |
| Anorectal Malformations | Not a EUROCAT subgroup | | Diagnosis codes in hospital or death record, and/or repair procedure code. See Ford et al., 2022[3] |
| Hirschrung's Disease | Q431 | | |
| Gastroschisis | Q793 | | |
| Unilateral renal agenesis | Q600 | | |
| Congenital hydronephrosis | Q620 | | |
| Hypospadias | Q54 | Q544 | Procedure code for primary repair of hypospadias (M731) with or without a diagnostic code, excluding concomitant codes Q560-Q564, E250, E258, E259, E345, Q640, Q641. See Wilkinson et al.,2017[4] |
| Club foot – talipes equinovarus | Q660 | | |
| Polydactyly | Q69 | | |
| Syndactyly | Q70 | | |
| Craniosynostosis | Q750 | | |
| **Chromosomal anomalies** | | | |
| Down syndrome | Q90 | | |
| Turner syndrome | Q96 | | |
| Klinefelter syndrome | Q980-Q984 | | |
| Di George syndrome | D821 | | |
| Karyotype XXX (Trisomy X) | Q970 | | |

*CHD: congenital heart defect. GA: gestational age. HES: Hospital Episode Statistics. PDA: patent ductus arteriosus.*

[a] Cases with more than one anomaly are only counted once

[b] Included in Severe CHD subgroup

[c] For each individual subgroup (excepting Any CA and Chromosomal anomalies), the primary analysis will be on isolated cases only (excluding those with chromosomal, genetic and multiple organ anomalies).

[d] Three-character codes indicate a range of codes where the fourth character can take any value (if defined); i.e. Q05 means Q051, Q052 through to Q059.

#### eTable2: Summary of Educational Outcomes

| National Curriculum (NC) Year | Typical pupil age, years | Key Stage (KS) | Academic Years analysed | Attainment measures |
|---|---|---|---|---|
| Reception | 4/5 | Early Years Foundation Stage Profile (EYFSP) | 2008/09 to 2011/12 | 13 Areas of Learning (scales), each scored 0-9 (highest); 6+ is considered expected<br>1. Good Level of Development (GLD)* = achieved 78 or more scale points across all 13 scales AND 6 or more scale points in each of the 3 scales of Personal, Social and Emotional Development (PSE) and 4 scales of Communication, Language and Literacy (CLL).<br>2. Expected English = 24+ total CLL scale points<br>3. Expected Maths = 18+ total scale points for Problem Solving, Reasoning and Numeracy (PSRN), comprising 3 scales<br>4. Standardised score English = total CLL scale points<br>5. Standardised score Maths = total PSRN scale points |
| | | | 2012/13 | 17 Early Learning Goals (ELG), each scored from 0-3 (highest); 2+ considered expected.<br>1. Good Level of Development (GLD) = achieved 2 or 3 in each of 12 ELG from Communication and Language (COM), Physical Development (PHY), Personal, Social and Emotional Development (PSE), Literacy (LIT) and Mathematics (MAT)<br>2. Expected English = 2+ in each of COM_G01, COM_G02, COM_G03 and LIT_G09, LIT_G10<br>3. Expected Maths = 2+ in each of MAT_G11, MAT_G12<br>4. Standardised score English = combined score across COM and LIT ELGs<br>5. Standardised score Maths = combined score across MAT ELGs |
| Year 2 | 6/7 | KS1 | 2010/11 to 2014/15 | Awarded levels based on teacher-assessment (<1/1/2C/2B/2A/3/4/5/6); levels were converted to point scores (<1=3, 1=9, 2C=13, 2B=15, 2A=17, 3=21, 4=27, 5=33, 6=39)<br>1. Expected English: Awarded level 2B and above in both Reading and Writing<br>2. Expected Maths: Awarded level 2B and above in Maths<br>3. Point Score in English (Reading and Writing combined)<br>4. Point Score in Maths |
| Year 6 | 10/11 | KS2 | 2014/15 | 1. Expected English: achieved level 4 or above in Reading<br>2. Expected Maths: achieved level 4 or above in Maths<br>3. Total test marks for Reading (0-50)<br>4. Total test marks for Maths (0-100) |
| | | | 2015-16 to 2018/19 | 1. Reached expected standard in Reading test<br>2. Reached expected standard in Maths test<br>3. Total test marks for Reading (0-50)<br>4. Total test marks for Maths (0-110) |

*CLL: Communication, Language and Literacy; ELG: Early Learning Goals; EYFSP: Early Years Foundation Stage Profile; GLD: Good Level of Development; KS: Key Stage; NC: National Curriculum; PSE: Personal, Social and Emotional Development; PSRN: Problem Solving, Reasoning and Numeracy*
* GLD was formally defined only for 2011/12; we applied the same formula to derive GLD for years prior

#### eTable3: Number of children enrolled in primary school and deaths, by age and congenital anomaly (CA) status

| Group | In Reception census (age 5) N (%) | In Year 2 census (age 7) N (%) | In Year 6 census (age 11) N (%) | Died during primary school N (%) |
|---|---|---|---|---|
| Children without CA | 2,272,742 (100.0) | 2,241,658 (98.6) | 2,181,797 (96.0) | 899 (0.04) |
| Children with any CA | 78,847 (100.0) | 77,802 (98.7) | 75,748 (96.1) | 330 (0.4) |

*CA: congenital anomaly*

eTable4: Number (%) of children reaching a Good Level of Development (GLD) at EYFSP, by congenital anomaly (CA) subgroup. Subgroups include only individuals with isolated CAs (except for Any CA and chromosomal anomalies).

| | **Male** | | **Female** | | **Total** | |
|---|---|---|---|---|---|---|
| **Subgroup** | N enrolled | N (%) reached GLD | N enrolled | N (%) reached GLD | N enrolled | N (%) reached GLD |
| No CA | 1,160,586 | 558,684 (48.1) | 1,112,156 | 736,765 (66.2) | 2,272,742 | 1,295,449 (57.0) |
| **Any CA** | 47,101 | 18,741 (39.8) | 31,746 | 17,313 (54.5) | 78,847 | 36,054 (45.7) |
| Neural Tube Defects | 128 | 41 (32.0) | 147 | 71 (48.3) | 275 | 112 (40.7) |
| Hydrocephalus | 242 | 40 (16.5) | 148 | 46 (31.1) | 390 | 86 (22.1) |
| Congenital Cataract | 181 | 67 (37.0) | 201 | 109 (54.2) | 382 | 176 (46.1) |
| Congenital Heart Defects (CHD) | 6,086 | 2,206 (36.2) | 5,282 | 2,720 (51.5) | 11,368 | 4,926 (43.3) |
| Ventricular Septal Defect* | 1,316 | 556 (42.2) | 1,315 | 750 (57.0) | 2,631 | 1,306 (49.6) |
| Pulmonary Valve Stenosis* | 210 | 74 (35.2) | 219 | 111 (50.7) | 429 | 185 (43.1) |
| PDA as only CHD in term infants | 1,071 | 313 (29.2) | 997 | 487 (48.8) | 2,068 | 800 (38.7) |
| Severe CHD | 1,805 | 682 (37.8) | 1,186 | 592 (49.9) | 2,991 | 1,274 (42.6) |
| Atrioventricular Septal Defect | 161 | 54 (33.5) | 176 | 80 (45.5) | 337 | 134 (39.8) |
| Tetralogy of Fallot | 313 | 117 (37.4) | 215 | 106 (49.3) | 528 | 223 (42.2) |
| Hypoplastic Left Heart | 126 | 35 (27.8) | 87 | 30 (34.5) | 213 | 65 (30.5) |
| Respiratory | 413 | 140 (33.9) | 321 | 166 (51.7) | 734 | 306 (41.7) |
| Cleft Lip | 567 | 248 (43.7) | 287 | 170 (59.2) | 854 | 418 (48.9) |
| Cleft Palate | 491 | 175 (35.6) | 543 | 275 (50.6) | 1,034 | 450 (43.5) |
| Cleft Lip and Palate | 711 | 254 (35.7) | 375 | 202 (53.9) | 1,086 | 456 (42.0) |
| Digestive System | 1,656 | 704 (42.5) | 1,351 | 782 (57.9) | 3,007 | 1,486 (49.4) |
| Anorectal Malformations | 103 | 40 (38.8) | 96 | 54 (56.2) | 199 | 94 (47.2) |
| Hirschsprung's Disease | 339 | 154 (45.4) | 115 | 65 (56.5) | 454 | 219 (48.2) |
| Gastroschisis | 395 | 142 (36.0) | 404 | 229 (56.7) | 799 | 371 (46.4) |
| Unilateral Renal Agenesis | 215 | 99 (46.0) | 108 | 66 (61.1) | 323 | 165 (51.1) |
| Congenital Hydronephrosis | 2,614 | 1,228 (47.0) | 918 | 606 (66.0) | 3,532 | 1,834 (51.9) |
| Hypospadias | 3,302 | 1,455 (44.1) | -- | -- | 3,302 | 1,455 (44.1) |
| Club Foot - Talipes Equinovarus | 1,280 | 575 (44.9) | 933 | 573 (61.4) | 2,213 | 1,148 (51.9) |
| Polydactyly | 2,004 | 880 (43.9) | 1,476 | 931 (63.1) | 3,480 | 1,811 (52.0) |
| Syndactyly | 1,012 | 437 (43.2) | 533 | 330 (61.9) | 1,545 | 767 (49.6) |
| Craniosynostosis | 349 | 143 (41.0) | 177 | 107 (60.5) | 526 | 250 (47.5) |

| | | | | | | |
|---|---|---|---|---|---|---|
| Down Syndrome | 1,184 | † | 950 | 16 (1.7) | 2,134 | † |
| Turner Syndrome | -- | -- | 115 | 37 (32.2) | 115 | 37 (32.2) |
| Klinefelter Syndrome | 72 | 23 (31.9) | -- | -- | 72 | 23 (31.9) |
| Di George Syndrome | 67 | † | 69 | † | 136 | † |
| Karyotype XXX | -- | -- | 40 | 22 (55.0) | 40 | 22 (55.0) |

*CA=congenital anomaly; CHD=congenital heart defect; ; EYFSP= Early Years Foundation Stage Profile; GLD=Good Level of Development; PDA=patent ductus arteriosus*

*without co-occurring severe CHD

-- not applicable

† suppressed count (<10) or dependent quantity

#### eTable5: Comparison of outcomes from congenital anomaly subgroups defined by EUROCAT and an alternative (Other) codelist

*birth prevalence based on all cases; number enrolled includes isolated cases only*

| | EYFSP | | KS1 | | KS2 | |
|---|---|---|---|---|---|---|
| | EUROCAT | Other[1] | EUROCAT | Other[1] | EUROCAT | Other[1] |
| **Severe CHD** | | | | | | |
| birth prevalence (per 10,000) | 21.8 | 37.3 | -- | -- | -- | -- |
| number enrolled | 2,991 | 4,785 | 2,946 | 4,721 | 2,854 | 4,580 |
| % achieved GLD | 42.6 | 42.2 | -- | -- | -- | -- |
| % achieved expected English | 54.6 | 53.5 | 51.4 | 51.5 | 60.4 | 60.5 |
| % achieved expected Maths | 66.2 | 64.6 | 63.4 | 62.2 | 62.9 | 60.9 |
| **Hypospadias** | | | | | | |
| birth prevalence (per 10,000) | 28.2 | 16.8 | | | | |
| number enrolled | 5,686 | 3,302 | 5,623 | 3,265 | 5,493 | 3,195 |
| % achieved GLD | 43.6 | 44.1 | -- | -- | -- | -- |
| % achieved expected English | 57.0 | 57.8 | 55.5 | 55.5 | 68.8 | 69.5 |
| % achieved expected Maths | 71.4 | 71.8 | 73.1 | 73.8 | 73.5 | 74.2 |
| **Cleft Palate** | | | | | | |
| birth prevalence (per 10,000) | 6.8 | 6.3 | -- | -- | -- | -- |
| number enrolled | 956 | 1,034 | 943 | 1,023 | 924 | 1,001 |
| % achieved GLD | 43.7 | 43.5 | -- | -- | -- | -- |
| % achieved expected English | 54.3 | 54.6 | 53.0 | 53.3 | 65.4 | 65.3 |
| % achieved expected Maths | 65.6 | 66.1 | 63.9 | 64.5 | 65.6 | 65.4 |
| **Cleft lip with or without palate[2]** | | | | | | |
| birth prevalence (per 10,000) | 8.8 | 7.8 | -- | -- | -- | -- |
| number enrolled | 1,748 | 1,687 | 1,724 | 1,663 | 1,683 | 1,623 |
| % achieved GLD | 45.0 | 45.1 | -- | -- | -- | -- |
| % achieved expected English | 57.6 | 57.7 | 55.8 | 55.7 | 67.0 | 67.0 |
| % achieved expected Maths | 70.1 | 70.3 | 69.4 | 69.3 | 70.1 | 70.1 |
| **Cleft, any** | | | | | | |
| birth prevalence (per 10,000) | 15.6 | 12.8 | -- | -- | -- | -- |
| number enrolled | 2,704 | 2,480 | 2,667 | 2,448 | 2,607 | 2,391 |
| % achieved GLD | 44.5 | 44.6 | -- | -- | -- | -- |
| % achieved expected English | 56.4 | 56.5 | 54.8 | 54.7 | 66.4 | 66.5 |
| % achieved expected Maths | 68.5 | 69.2 | 67.5 | 67.8 | 68.5 | 68.6 |

*CHD=congenital heart defect; GLD=Good Level of Development; EYFSP=Early Years Foundation Stage Profile; KS1=Key Stage 1; KS2=Key Stage 2*

[1] Severe CHD: Gimeno et al. (2023); Hypospadias: Wilkinson et al. (2017); Clefts (all): Fitzsimons et al. (2018)

[2] Other category comprises individuals with a Q36 or Q37 ICD-10 diagnosis code

-- not applicable

#### eTable6: Percentage of children with structural congenital anomalies (CAs) achieving expected level of attainment for English and Maths, by key stage and malformation type

| | English | | | | | | | | | Maths | | | | | | | | |
|---|---|---|---|---|---|---|---|---|---|---|---|---|---|---|---|---|---|---|
| | EYFSP | | | KS1 | | | KS2 | | | EYFSP | | | KS1 | | | KS2 | | |
| **CA subgroup** | Isolated | Multiple | Genetic | Isolated | Multiple | Genetic | Isolated | Multiple | Genetic | Isolated | Multiple | Genetic | Isolated | Multiple | Genetic | Isolated | Multiple | Genetic |
| Neural Tube Defects | 50.2 | 35.0 | -- | 54.0 | 32.4 | -- | 59.8 | 45.1 | -- | 63.3 | 47.5 | -- | 59.9 | 39.0 | -- | 57.9 | 41.3 | -- |
| Hydrocephalus | 31.0 | 24.4 | 0.0 | 29.3 | 22.2 | 0.0 | 36.9 | 34.2 | 0.0 | 39.5 | 30.2 | 0.0 | 36.8 | 24.8 | 0.0 | 35.6 | 28.8 | 0.0 |
| Congenital Cataract | 58.9 | 0.0 | 0.0 | 57.6 | 0.0 | 0.0 | 67.5 | 20.8 | 0.0 | 70.9 | 38.9 | 0.0 | 69.2 | 0.0 | 0.0 | 67.2 | 39.6 | 0.0 |
| Congenital Heart Defects (CHD) | 54.2 | 40.5 | 12.3 | 52.3 | 38.6 | 9.7 | 62.9 | 48.4 | 13.3 | 65.3 | 49.8 | 17.1 | 63.3 | 46.4 | 12.0 | 62.6 | 46.9 | 12.1 |
| Ventricular Septal Defect* | 59.9 | 40.9 | 9.9 | 57.7 | 39.1 | 7.4 | 68.4 | 47.7 | 12.6 | 70.7 | 51.1 | 15.2 | 68.7 | 48.2 | 9.6 | 68.3 | 49.7 | 11.2 |
| Pulmonary Valve Stenosis* | 53.8 | 33.8 | 19.2 | 55.0 | 15.6 | 0.0 | 64.4 | 44.3 | 0.0 | 66.0 | 44.6 | 25.0 | 64.5 | 39.1 | 0.0 | 60.8 | 41.0 | 0.0 |
| PDA as only CHD in term infants | 49.4 | 38.0 | 13.3 | 47.8 | 36.0 | 11.2 | 61.1 | 46.6 | 21.2 | 60.7 | 48.5 | 25.2 | 57.4 | 45.4 | 20.3 | 56.5 | 44.8 | 18.2 |
| Severe CHD | 54.6 | 42.0 | 9.8 | 51.4 | 36.5 | 7.4 | 60.4 | 47.4 | 8.9 | 66.2 | 50.3 | 12.6 | 63.4 | 43.9 | 8.7 | 62.9 | 46.4 | 7.9 |
| Atrioventricular Septal Defect | 54.6 | 29.2 | 0.0 | 52.0 | 15.8 | 0.0 | 55.7 | 35.6 | 0.0 | 65.0 | 38.5 | 3.0 | 61.7 | 31.6 | 0.0 | 58.2 | 28.7 | 0.0 |
| Tetralogy of Fallot | 55.1 | 43.9 | 10.0 | 51.8 | 38.5 | 6.8 | 61.3 | 48.9 | 7.5 | 67.4 | 51.4 | 19.3 | 62.9 | 46.2 | 13.5 | 65.7 | 48.9 | 6.8 |
| Hypoplastic Left Heart | 41.3 | 24.4 | -- | 39.9 | 0.0 | -- | 51.5 | 0.0 | -- | 56.3 | 24.4 | -- | 51.0 | 0.0 | -- | 47.4 | 0.0 | -- |
| Respiratory | 52.2 | 41.4 | 21.7 | 52.3 | 40.6 | 0.0 | 62.2 | 52.4 | 27.1 | 63.6 | 52.7 | 27.0 | 63.2 | 49.4 | 21.2 | 63.2 | 53.9 | 26.2 |
| Cleft Lip | 61.4 | 53.3 | 0.0 | 61.6 | 49.5 | 0.0 | 70.0 | 62.7 | 0.0 | 73.5 | 60.0 | 0.0 | 72.8 | 64.1 | 0.0 | 75.3 | 60.8 | 0.0 |
| Cleft Palate | 54.6 | 37.2 | 42.6 | 53.3 | 35.6 | 41.2 | 65.3 | 41.5 | 49.1 | 66.1 | 43.9 | 51.8 | 64.5 | 45.4 | 49.8 | 65.4 | 44.1 | 51.6 |
| Cleft Lip and Palate | 54.3 | 40.8 | 38.4 | 52.3 | 43.6 | 41.2 | 64.6 | 55.6 | 55.6 | 66.9 | 52.4 | 48.8 | 67.3 | 52.9 | 45.9 | 66.6 | 54.1 | 46.9 |
| Digestive System | 60.2 | 51.6 | 25.4 | 59.4 | 51.2 | 25.1 | 69.8 | 61.3 | 29.6 | 72.0 | 62.8 | 33.0 | 72.1 | 60.3 | 27.3 | 71.3 | 60.8 | 29.1 |
| Anorectal Malformations | 57.8 | 51.2 | 40.2 | 59.2 | 54.0 | 39.2 | 70.8 | 62.8 | 47.9 | 68.8 | 63.0 | 54.1 | 69.9 | 62.8 | 44.2 | 69.3 | 64.8 | 44.5 |
| Hirschsprung's Disease | 59.7 | 57.6 | 0.0 | 58.0 | 58.2 | 0.0 | 69.6 | 65.1 | 0.0 | 73.6 | 69.6 | 0.0 | 72.8 | 69.2 | 0.0 | 71.9 | 66.3 | 0.0 |
| Gastroschisis | 57.6 | 51.9 | -- | 54.2 | 43.7 | -- | 65.6 | 65.6 | -- | 68.1 | 63.9 | -- | 66.7 | 59.5 | -- | 67.0 | 62.3 | -- |
| Unilateral Renal Agenesis | 61.6 | 37.3 | 0.0 | 66.5 | 36.6 | 0.0 | 66.3 | 52.1 | 0.0 | 73.7 | 54.9 | 0.0 | 77.2 | 47.5 | 0.0 | 72.8 | 51.0 | 0.0 |

| | | | | | | | | | | | | | | | | | | |
|---|---|---|---|---|---|---|---|---|---|---|---|---|---|---|---|---|---|---|
| Congenital Hydronephrosis | 64.2 | 46.4 | 20.6 | 61.3 | 45.6 | 9.5 | 74.1 | 55.5 | 29.8 | 75.5 | 60.5 | 29.9 | 77.5 | 56.2 | 24.8 | 76.4 | 55.8 | 26.9 |
| Hypospadias | 57.8 | 47.8 | 55.1 | 55.5 | 43.3 | 53.8 | 69.5 | 57.5 | 68.2 | 71.8 | 60.1 | 70.8 | 73.8 | 60.6 | 71.4 | 74.2 | 60.8 | 72.2 |
| Club Foot - Talipes Equinovarus | 63.4 | 43.1 | 31.8 | 61.0 | 44.3 | 32.1 | 71.1 | 57.8 | 41.3 | 74.3 | 53.6 | 45.9 | 73.9 | 52.3 | 42.9 | 73.0 | 56.6 | 38.8 |
| Polydactyly | 63.0 | 47.1 | 57.6 | 63.2 | 44.1 | 55.7 | 72.5 | 55.5 | 63.3 | 74.3 | 58.0 | 65.5 | 74.2 | 52.8 | 64.5 | 74.1 | 56.3 | 64.8 |
| Syndactyly | 64.3 | 54.4 | 28.6 | 62.3 | 54.8 | 32.4 | 72.5 | 65.6 | 42.1 | 75.2 | 65.9 | 43.8 | 75.4 | 65.3 | 39.6 | 74.0 | 65.2 | 37.4 |
| Craniosynostosis | 60.5 | 38.1 | 29.8 | 57.6 | 39.0 | 25.7 | 69.1 | 48.7 | 29.7 | 73.0 | 53.2 | 36.8 | 69.4 | 46.3 | 31.0 | 68.5 | 48.7 | 36.0 |

*CA=congenital anomalies; CHD=congenital heart defect; EYFSP=Early Years Foundation Stage Profile; KS1=Key Stage 1; KS2=Key Stage 2; PDA=patent ductus arteriosus*

*without co-occurring severe CHD

-- no observations

Genetic category includes chromosomal, genetic and non-system specific anomalies eTable7: Adjusted risk ratios for achieving expected levels of attainment in English, comparing children with congenital anomalies (CAs) and children without, by key stage and CA subgroup, for subset of children assessed at three key stages. For each comparison, number of children without CAs= 2,008,951

| | | EYFSP | | KS1 | | KS2 | |
|---|---|---|---|---|---|---|---|
| **subgroup** | **Cases, N** | RR (95%CI) Sex adj. | RR (95%CI) Full adj. | RR (95%CI) Sex adj. | RR (95%CI) Full adj. | RR (95%CI) Sex adj. | RR (95%CI) Full adj. |
| Any CA | 62,778 | 0.92 (0.92,0.93) | 0.93 (0.92,0.93) | 0.93 (0.92,0.93) | 0.93 (0.92,0.94) | 0.95 (0.94,0.95) | 0.95 (0.95,0.95) |
| Neural Tube Defects | 201 | 0.83 (0.74,0.93) | 0.86 (0.77,0.97) | 0.90 (0.81,1.00) | 0.94 (0.84,1.04) | 0.91 (0.83,0.99) | 0.93 (0.86,1.01) |
| Hydrocephalus | 211 | 0.70 (0.61,0.80) | 0.72 (0.63,0.83) | 0.74 (0.65,0.84) | 0.76 (0.67,0.87) | 0.80 (0.72,0.88) | 0.81 (0.73,0.90) |
| Congenital Cataract | 308 | 0.88 (0.81,0.96) | 0.90 (0.83,0.97) | 0.90 (0.83,0.98) | 0.91 (0.84,0.99) | 0.96 (0.90,1.02) | 0.96 (0.90,1.02) |
| Congenital Heart Defects (CHD) | 8,731 | 0.86 (0.85,0.88) | 0.87 (0.86,0.89) | 0.86 (0.84,0.87) | 0.87 (0.85,0.88) | 0.90 (0.88,0.91) | 0.90 (0.89,0.92) |
| Ventricular Septal Defect | 2,202 | 0.92 (0.89,0.95) | 0.93 (0.90,0.95) | 0.91 (0.88,0.94) | 0.92 (0.89,0.95) | 0.95 (0.93,0.97) | 0.96 (0.93,0.98) |
| Pulmonary Valve Stenosis | 344 | 0.82 (0.75,0.89) | 0.82 (0.75,0.89) | 0.87 (0.80,0.95) | 0.88 (0.81,0.95) | 0.91 (0.85,0.97) | 0.91 (0.85,0.97) |
| PDA as only CHD in term infants | 1,359 | 0.81 (0.77,0.85) | 0.83 (0.79,0.86) | 0.81 (0.78,0.85) | 0.83 (0.79,0.87) | 0.89 (0.86,0.92) | 0.90 (0.87,0.93) |
| Severe CHD | 2,311 | 0.88 (0.85,0.91) | 0.89 (0.86,0.92) | 0.87 (0.84,0.90) | 0.87 (0.84,0.90) | 0.87 (0.85,0.90) | 0.88 (0.85,0.90) |
| Atrioventricular Septal Defect | 268 | 0.87 (0.79,0.95) | 0.88 (0.81,0.97) | 0.86 (0.78,0.94) | 0.87 (0.79,0.96) | 0.82 (0.75,0.89) | 0.83 (0.76,0.90) |
| Tetralogy of Fallot | 423 | 0.89 (0.83,0.96) | 0.90 (0.84,0.97) | 0.86 (0.80,0.94) | 0.87 (0.80,0.94) | 0.89 (0.83,0.94) | 0.89 (0.84,0.95) |
| Hypoplastic Left Heart | 145 | 0.73 (0.62,0.85) | 0.75 (0.64,0.88) | 0.73 (0.62,0.86) | 0.76 (0.65,0.89) | 0.80 (0.71,0.91) | 0.82 (0.72,0.92) |
| Respiratory | 555 | 0.84 (0.79,0.90) | 0.85 (0.79,0.91) | 0.87 (0.81,0.93) | 0.88 (0.82,0.94) | 0.90 (0.85,0.95) | 0.90 (0.86,0.95) |
| Cleft Lip | 740 | 0.94 (0.89,0.99) | 0.95 (0.90,1.00) | 0.98 (0.93,1.03) | 0.99 (0.94,1.04) | 0.96 (0.92,1.00) | 0.96 (0.92,1.00) |
| Cleft Palate | 877 | 0.83 (0.79,0.88) | 0.84 (0.79,0.88) | 0.82 (0.78,0.87) | 0.83 (0.79,0.88) | 0.91 (0.87,0.95) | 0.91 (0.88,0.95) |
| Cleft Lip and Palate | 932 | 0.84 (0.79,0.88) | 0.84 (0.80,0.89) | 0.84 (0.80,0.89) | 0.86 (0.81,0.91) | 0.89 (0.86,0.93) | 0.90 (0.87,0.94) |
| Digestive System | 2,476 | 0.91 (0.88,0.93) | 0.91 (0.89,0.94) | 0.92 (0.90,0.95) | 0.93 (0.90,0.96) | 0.95 (0.93,0.97) | 0.95 (0.93,0.98) |
| Anorectal Malformations | 152 | 0.88 (0.78,1.00) | 0.89 (0.79,1.01) | 0.99 (0.89,1.11) | 1.01 (0.91,1.12) | 1.00 (0.92,1.08) | 1.01 (0.93,1.09) |
| Hirschsprung's Disease | 369 | 0.95 (0.88,1.02) | 0.96 (0.89,1.03) | 0.93 (0.86,1.01) | 0.94 (0.87,1.02) | 0.96 (0.90,1.02) | 0.97 (0.91,1.03) |
| Gastroschisis | 657 | 0.83 (0.78,0.89) | 0.92 (0.87,0.98) | 0.81 (0.76,0.87) | 0.92 (0.86,0.99) | 0.86 (0.81,0.90) | 0.93 (0.88,0.98) |
| Unilateral Renal Agenesis | 268 | 0.98 (0.90,1.06) | 0.98 (0.91,1.07) | 1.07 (0.99,1.16) | 1.08 (1.00,1.16) | 0.91 (0.85,0.99) | 0.92 (0.85,0.99) |
| Congenital Hydronephrosis | 3,087 | 0.99 (0.96,1.01) | 0.98 (0.95,1.00) | 0.98 (0.95,1.00) | 0.97 (0.94,0.99) | 1.00 (0.98,1.02) | 1.00 (0.98,1.02) |
| Hypospadias | 2,866 | 0.96 (0.93,0.99) | 0.96 (0.93,0.99) | 0.96 (0.93,0.99) | 0.96 (0.93,0.99) | 0.97 (0.95,1.00) | 0.98 (0.96,1.00) |
| Club Foot - Talipes Equinovarus | 1,919 | 0.97 (0.94,1.00) | 0.97 (0.94,1.00) | 0.95 (0.92,0.98) | 0.95 (0.92,0.98) | 0.96 (0.94,0.99) | 0.97 (0.94,0.99) |
| Polydactyly | 3,017 | 0.95 (0.93,0.98) | 0.98 (0.96,1.00) | 0.98 (0.95,1.00) | 0.99 (0.96,1.01) | 0.98 (0.96,1.00) | 0.98 (0.97,1.00) |

| | | | | | | | |
|---|---|---|---|---|---|---|---|
| Syndactyly | 1,364 | 0.97 (0.93,1.01) | 0.95 (0.92,0.99) | 0.98 (0.94,1.02) | 0.97 (0.94,1.01) | 0.99 (0.96,1.01) | 0.98 (0.95,1.01) |
| Craniosynostosis | 435 | 0.95 (0.89,1.02) | 0.94 (0.88,1.00) | 0.92 (0.86,0.99) | 0.91 (0.85,0.98) | 0.97 (0.92,1.03) | 0.97 (0.92,1.02) |
| Down Syndrome | 58 | 0.40 (0.26,0.59) | 0.39 (0.26,0.58) | 0.38 (0.25,0.57) | 0.37 (0.24,0.56) | 0.52 (0.38,0.70) | 0.50 (0.37,0.69) |
| Turner Syndrome | 93 | 0.62 (0.50,0.77) | 0.62 (0.50,0.76) | 0.67 (0.55,0.82) | 0.67 (0.55,0.82) | 0.80 (0.69,0.93) | 0.79 (0.68,0.92) |
| Klinefelter Syndrome | 49 | 0.83 (0.64,1.08) | 0.79 (0.61,1.02) | 0.53 (0.35,0.79) | 0.50 (0.33,0.75) | 0.49 (0.34,0.70) | 0.46 (0.32,0.67) |
| Di George Syndrome | 46 | 0.39 (0.25,0.62) | 0.39 (0.25,0.62) | 0.25 (0.13,0.46) | 0.24 (0.13,0.46) | 0.30 (0.18,0.51) | 0.30 (0.18,0.50) |
| Karyotype XXX | 32 | 0.89 (0.70,1.12) | 0.81 (0.64,1.03) | 0.41 (0.24,0.68) | 0.38 (0.22,0.63) | 0.50 (0.33,0.76) | 0.46 (0.30,0.71) |

*CI=confidence interval; EYFSP=Early Years Foundation Stage Profile; KS1=Key Stage 1; KS2=Key Stage 2; RR=risk ratio; Sex adj.= adjusted for sex at birth; Full adj.=adjusted for sex, maternal age at birth, ethnicity, income deprivation affecting children index (IDACI) quintile, free school meals eligibility (FSME)* eTable8: Adjusted risk ratios for achieving expected levels of attainment in Maths, comparing children with congenital anomalies (CAs) and children without, by key stage and CA subgroup, for subset of children assessed at three key stages. For each comparison, number of children without CAs= 2,009,287

| | | EYFSP | | KS1 | | KS2 | |
|---|---|---|---|---|---|---|---|
| **subgroup** | **Cases, N** | RR (95%CI) Sex adj. | RR (95%CI) Full adj. | RR (95%CI) Sex adj. | RR (95%CI) Full adj. | RR (95%CI) Sex adj. | RR (95%CI) Full adj. |
| Any CA | 62,725 | 0.95 (0.94,0.95) | 0.95 (0.95,0.95) | 0.93 (0.93,0.94) | 0.94 (0.93,0.94) | 0.94 (0.93,0.94) | 0.94 (0.94,0.94) |
| Neural Tube Defects | 198 | 0.89 (0.81,0.97) | 0.91 (0.84,0.99) | 0.88 (0.80,0.96) | 0.90 (0.82,0.98) | 0.88 (0.80,0.96) | 0.89 (0.82,0.98) |
| Hydrocephalus | 211 | 0.78 (0.70,0.86) | 0.79 (0.71,0.88) | 0.77 (0.69,0.85) | 0.78 (0.70,0.87) | 0.74 (0.66,0.83) | 0.75 (0.67,0.84) |
| Congenital Cataract | 312 | 0.95 (0.89,1.01) | 0.96 (0.90,1.02) | 0.93 (0.88,0.99) | 0.94 (0.89,1.00) | 0.92 (0.87,0.99) | 0.93 (0.87,0.99) |
| Congenital Heart Defects (CHD) | 8,705 | 0.90 (0.89,0.92) | 0.91 (0.90,0.92) | 0.88 (0.87,0.89) | 0.88 (0.87,0.90) | 0.88 (0.87,0.90) | 0.89 (0.88,0.90) |
| Ventricular Septal Defect | 2,194 | 0.95 (0.93,0.97) | 0.96 (0.93,0.98) | 0.93 (0.90,0.95) | 0.93 (0.91,0.95) | 0.93 (0.91,0.96) | 0.94 (0.92,0.96) |
| Pulmonary Valve Stenosis | 345 | 0.88 (0.83,0.94) | 0.88 (0.83,0.94) | 0.86 (0.80,0.92) | 0.86 (0.81,0.93) | 0.82 (0.76,0.89) | 0.82 (0.76,0.89) |
| PDA as only CHD in term infants | 1,349 | 0.86 (0.83,0.89) | 0.87 (0.84,0.91) | 0.83 (0.80,0.87) | 0.84 (0.81,0.87) | 0.84 (0.80,0.87) | 0.84 (0.81,0.87) |
| Severe CHD | 2,312 | 0.92 (0.90,0.94) | 0.93 (0.90,0.95) | 0.88 (0.86,0.91) | 0.89 (0.86,0.91) | 0.89 (0.87,0.91) | 0.89 (0.87,0.91) |
| Atrioventricular Septal Defect | 266 | 0.91 (0.85,0.98) | 0.92 (0.86,0.99) | 0.88 (0.82,0.95) | 0.89 (0.83,0.96) | 0.84 (0.77,0.91) | 0.85 (0.78,0.92) |
| Tetralogy of Fallot | 427 | 0.92 (0.87,0.98) | 0.93 (0.88,0.98) | 0.86 (0.81,0.92) | 0.87 (0.82,0.92) | 0.92 (0.87,0.97) | 0.92 (0.87,0.97) |
| Hypoplastic Left Heart | 141 | 0.83 (0.74,0.93) | 0.85 (0.76,0.95) | 0.80 (0.70,0.90) | 0.81 (0.72,0.91) | 0.77 (0.68,0.88) | 0.78 (0.69,0.89) |
| Respiratory | 556 | 0.88 (0.84,0.93) | 0.89 (0.84,0.93) | 0.87 (0.83,0.92) | 0.88 (0.83,0.93) | 0.89 (0.84,0.94) | 0.89 (0.85,0.94) |
| Cleft Lip | 746 | 0.96 (0.92,1.00) | 0.96 (0.92,1.00) | 0.94 (0.90,0.98) | 0.95 (0.91,0.98) | 0.99 (0.95,1.02) | 1.00 (0.96,1.03) |
| Cleft Palate | 887 | 0.88 (0.84,0.91) | 0.88 (0.84,0.91) | 0.85 (0.82,0.89) | 0.86 (0.82,0.89) | 0.89 (0.85,0.93) | 0.89 (0.86,0.93) |
| Cleft Lip and Palate | 943 | 0.88 (0.84,0.92) | 0.89 (0.85,0.92) | 0.89 (0.86,0.93) | 0.90 (0.86,0.93) | 0.89 (0.85,0.92) | 0.90 (0.86,0.94) |
| Digestive System | 2,470 | 0.95 (0.93,0.98) | 0.96 (0.94,0.98) | 0.94 (0.92,0.97) | 0.95 (0.93,0.97) | 0.95 (0.93,0.97) | 0.96 (0.94,0.98) |
| Anorectal Malformations | 155 | 0.93 (0.85,1.02) | 0.94 (0.86,1.02) | 0.94 (0.86,1.02) | 0.94 (0.86,1.03) | 0.93 (0.85,1.02) | 0.94 (0.86,1.03) |
| Hirschsprung's Disease | 366 | 0.98 (0.92,1.03) | 0.98 (0.93,1.03) | 0.96 (0.91,1.02) | 0.97 (0.91,1.02) | 0.96 (0.91,1.02) | 0.97 (0.92,1.02) |
| Gastroschisis | 657 | 0.88 (0.84,0.93) | 0.94 (0.90,0.99) | 0.85 (0.81,0.89) | 0.91 (0.87,0.96) | 0.86 (0.81,0.90) | 0.93 (0.88,0.98) |
| Unilateral Renal Agenesis | 269 | 0.99 (0.94,1.06) | 1.00 (0.94,1.06) | 1.02 (0.97,1.08) | 1.03 (0.97,1.08) | 0.96 (0.90,1.03) | 0.97 (0.91,1.03) |
| Congenital Hydronephrosis | 3,093 | 0.98 (0.97,1.00) | 0.98 (0.96,0.99) | 1.00 (0.99,1.02) | 1.00 (0.98,1.01) | 0.99 (0.97,1.01) | 0.99 (0.97,1.00) |
| Hypospadias | 2,871 | 0.97 (0.95,0.99) | 0.97 (0.95,0.99) | 0.97 (0.95,0.99) | 0.97 (0.95,0.99) | 0.98 (0.96,1.00) | 0.98 (0.96,1.00) |
| Club Foot - Talipes Equinovarus | 1,924 | 0.98 (0.96,1.00) | 0.98 (0.96,1.00) | 0.96 (0.94,0.99) | 0.96 (0.94,0.99) | 0.96 (0.94,0.99) | 0.97 (0.94,0.99) |
| Polydactyly | 3,017 | 0.97 (0.96,0.99) | 1.00 (0.98,1.02) | 0.97 (0.95,0.99) | 0.99 (0.97,1.00) | 0.97 (0.96,0.99) | 0.98 (0.96,1.00) |

| | | | | | | | |
|---|---|---|---|---|---|---|---|
| Syndactyly | 1,362 | 0.97 (0.94,1.00) | 0.96 (0.93,0.99) | 0.98 (0.95,1.00) | 0.97 (0.94,1.00) | 0.97 (0.95,1.00) | 0.97 (0.94,1.00) |
| Craniosynostosis | 434 | 0.99 (0.94,1.04) | 0.98 (0.93,1.02) | 0.93 (0.88,0.98) | 0.92 (0.88,0.97) | 0.93 (0.88,0.98) | 0.92 (0.88,0.97) |
| Down Syndrome | 53 | 0.55 (0.41,0.75) | 0.55 (0.41,0.74) | 0.42 (0.29,0.61) | 0.41 (0.28,0.60) | 0.47 (0.33,0.67) | 0.46 (0.33,0.65) |
| Turner Syndrome | 89 | 0.71 (0.59,0.84) | 0.70 (0.59,0.84) | 0.59 (0.48,0.73) | 0.59 (0.48,0.73) | 0.62 (0.50,0.76) | 0.62 (0.50,0.76) |
| Klinefelter Syndrome | 50 | 1.03 (0.89,1.18) | 0.98 (0.86,1.12) | 0.70 (0.55,0.89) | 0.67 (0.53,0.86) | 0.60 (0.45,0.80) | 0.58 (0.43,0.77) |
| Di George Syndrome | 44 | 0.37 (0.23,0.58) | 0.36 (0.23,0.57) | 0.31 (0.18,0.51) | 0.31 (0.18,0.51) | 0.28 (0.16,0.49) | 0.28 (0.16,0.49) |
| Karyotype XXX | 32 | 0.90 (0.74,1.10) | 0.84 (0.69,1.02) | 0.65 (0.47,0.90) | 0.61 (0.44,0.85) | 0.47 (0.30,0.74) | 0.45 (0.29,0.70) |

*CI=confidence interval; EYFSP=Early Years Foundation Stage Profile; KS1=Key Stage 1; KS2=Key Stage 2; RR=risk ratio; Sex adj.= adjusted for sex at birth; Full adj.=adjusted for sex, maternal age at birth, ethnicity, income deprivation affecting children index (IDACI) quintile, free school meals eligibility (FSME)*

#### eTable9: Distribution of subject standardised scores for selected congenital anomaly (CA) subgroups by key stage

| **English** | EYFSP | | KS1 | | KS2 | |
|---|---|---|---|---|---|---|
| **CA subgroup** | Number | mean (SD) | Number | mean (SD) | Number | mean (SD) |
| No CA | 2,255,528 | 0.03 (0.97) | 2,223,011 | 0.04 (0.92) | 2,121,289 | 0.02 (0.98) |
| Any CA | 77,829 | -0.33 (1.23) | 76,698 | -0.32 (1.20) | 67,152 | -0.12 (1.04) |
| Nervous System | 1,981 | -1.17 (1.58) | 1,957 | -1.16 (1.49) | 1,267 | -0.36 (1.11) |
| Congenital Heart Defects (CHD) | 11,240 | -0.36 (1.17) | 11,065 | -0.36 (1.14) | 9,813 | -0.22 (1.08) |
| Severe CHD | 2,963 | -0.37 (1.17) | 2,906 | -0.38 (1.16) | 2,531 | -0.27 (1.08) |
| Cleft Lip | 846 | -0.18 (1.06) | 831 | -0.11 (1.01) | 772 | -0.10 (1.02) |
| Cleft Palate | 1,030 | -0.37 (1.18) | 1,017 | -0.30 (1.16) | 915 | -0.16 (1.09) |
| Cleft Lip and Palate | 1,077 | -0.38 (1.11) | 1,066 | -0.30 (1.08) | 975 | -0.23 (1.07) |
| Digestive System | 2,983 | -0.19 (1.08) | 2,958 | -0.17 (1.05) | 2,712 | -0.10 (1.03) |
| Urinary System | 9,847 | -0.08 (1.02) | 9,683 | -0.08 (0.99) | 9,107 | -0.03 (1.00) |
| Congenital Hydronephrosis | 3,505 | -0.06 (1.02) | 3,441 | -0.08 (0.98) | 3,247 | -0.03 (0.99) |
| Hypospadias | 3,266 | -0.24 (1.03) | 3,239 | -0.22 (1.01) | 3,013 | -0.14 (1.01) |
| Polydactyly | 3,443 | -0.10 (1.04) | 3,401 | -0.07 (0.99) | 3,179 | -0.05 (0.97) |
| Down Syndrome | 2,007 | -2.75 (0.95) | 1,937 | -2.91 (0.60) | 66 | -1.40 (1.49) |
| Turner Syndrome | 132 | -0.86 (1.44) | 133 | -0.84 (1.38) | 101 | -0.20 (1.01) |
| Klinefelter Syndrome | 73 | -0.83 (1.21) | 73 | -1.04 (1.12) | 52 | -1.14 (1.11) |
| Di George Syndrome | 134 | -1.87 (1.06) | 130 | -2.03 (1.10) | 52 | -1.60 (1.02) |
| Karyotype XXX | 39 | -0.46 (1.06) | 39 | -0.63 (1.03) | 34 | -1.06 (1.08) |

| **Maths** | EYFSP | | KS1 | | KS2 | |
|---|---|---|---|---|---|---|
| **CA subgroup** | Number | mean (SD) | Number | mean (SD) | Number | mean (SD) |
| No CA | 2,255,528 | 0.03 (0.96) | 2,222,983 | 0.04 (0.96) | 2,122,249 | 0.01 (0.99) |
| Any CA | 77,829 | -0.33 (1.31) | 76,702 | -0.35 (1.30) | 67,133 | -0.15 (1.07) |
| Nervous System | 1,981 | -1.26 (1.79) | 1,957 | -1.33 (1.63) | 1,258 | -0.51 (1.19) |
| Congenital Heart Defects (CHD) | 11,240 | -0.36 (1.23) | 11,066 | -0.40 (1.22) | 9,796 | -0.29 (1.12) |
| Severe CHD | 2,963 | -0.35 (1.23) | 2,906 | -0.40 (1.23) | 2,539 | -0.26 (1.10) |
| Cleft Lip | 846 | -0.13 (1.09) | 831 | -0.09 (1.08) | 779 | -0.04 (1.02) |
| Cleft Palate | 1,030 | -0.34 (1.22) | 1,017 | -0.36 (1.21) | 926 | -0.27 (1.12) |
| Cleft Lip and Palate | 1,077 | -0.30 (1.14) | 1,066 | -0.28 (1.11) | 986 | -0.26 (1.11) |
| Digestive System | 2,983 | -0.19 (1.12) | 2,958 | -0.19 (1.12) | 2,709 | -0.12 (1.06) |
| Urinary System | 9,847 | -0.04 (1.04) | 9,683 | -0.02 (1.24) | 9,125 | -0.03 (1.04) |
| Congenital Hydronephrosis | 3,505 | 0.00 (1.02) | 3,441 | 0.00 (1.03) | 3,254 | 0.01 (1.01) |
| Hypospadias | 3,266 | -0.14 (1.08) | 3,239 | -0.11 (1.10) | 3,019 | -0.03 (1.04) |
| Polydactyly | 3,443 | -0.09 (1.07) | 3,401 | -0.11 (1.05) | 3,179 | -0.07 (1.01) |
| Down Syndrome | 2,007 | -3.12 (1.29) | 1,938 | -3.26 (0.74) | 60 | -1.31 (1.55) |
| Turner Syndrome | 132 | -0.96 (1.58) | 133 | -1.20 (1.40) | 97 | -0.75 (1.06) |
| Klinefelter Syndrome | 73 | -0.55 (1.29) | 73 | -0.90 (1.28) | 53 | -0.83 (1.15) |
| Di George Syndrome | 134 | -1.90 (1.30) | 130 | -2.23 (1.12) | 49 | -1.81 (1.02) |
| Karyotype XXX | 39 | -0.46 (1.19) | 39 | -0.84 (1.25) | 34 | -1.15 (1.09) |

*CA=congenital anomaly; CHD=congenital heart defect; EYFSP=Early Years Foundation Stage Profile; KS1=Key Stage 1; KS2=Key Stage 2; SD=standard deviation*
